# Postoperative Lymph is a Proximal Source of ctDNA for Detection of Recurrence in HPV-independent Head and Neck Cancer

**DOI:** 10.1101/2024.09.27.24314491

**Authors:** Seka Lazare, Zhuosheng Gu, Noah Earland, Adam Harmon, Maciej Pacula, Megan Rivera, Ashley Tellis, Damion Whitfield, Adam Benson, Sophie Gerndt, Peter Harris, Lucien Khalil, Ricardo Ramirez, Zhongping Xu, Benjamin Wahle, Sid Puram, Doug Adkins, Wade Thorstad, Daniel Zandberg, Rebecca Chernock, Heath Skinner, Matthew E Spector, Raja Seethala, Robert L. Ferris, Joshua D. Smith, Amy E. Greer, Ellen L. Verner, Andrew Georgiadis, Mark Sausen, Shakti H. Ramkissoon, Taylor J. Jensen, Marra S. Francis, Wendy Winckler, Aadel A. Chaudhuri, Jose P. Zevallos

## Abstract

**Purpose:** Relapse is a major cause of failure in human papillomavirus (HPV)-independent head and neck squamous cell carcinoma (HNSCC). Clinicopathologic criteria for adjuvant treatment remain imprecise and have not changed for decades. We investigated whether circulating tumor DNA (ctDNA) in lymphatic exudate collected via surgical drains (“lymph”) 24-hours after surgery identified molecular residual disease (MRD) and compared its performance to time-matched plasma.

**Experimental Design:** Using an ultra-sensitive tumor-informed sequencing approach, tumor variants were called in lymph and plasma to classify patients as ctDNA-positive or ctDNA-negative, trained in an initial cohort of 36 patients and replicated in an independent cohort of 37 patients. Progression-free survival (PFS) was compared in ctDNA+ vs. ctDNA-patients.

**Results:** Lymph identified MRD in two independent multi-site cohorts (initial cohort sensitivity = 76%, specificity = 63%, *P* = 0.01; replication cohort sensitivity = 65%, specificity = 70%, *P* = 0.04). Lymph performance was enhanced in locoregional relapse (sensitivity = 78%, specificity = 67%, *P* = 0.0004) and generalized to early-stage patients. Analysis of matched plasma collected at this early timepoint was not predictive of recurrence (sensitivity = 35%, specificity = 72%, *P* = 0.7). In patients with intermediate-risk pathology, lymph ctDNA was associated with recurrence (sensitivity = 88%, specificity = 67%, *P* = 0.0008), suggesting an opportunity for improved stratification of patients who may benefit from additional adjuvant treatment.

**Conclusion:** Postoperative lymph is a novel, proximal, and early source of MRD with the potential to introduce more precision into adjuvant therapy decision-making and improve outcomes, especially for intermediate-risk HPV-independent HNSCC patients.

**Translational Relevance:** Postoperative lymphatic exudate represents a proximal analyte for MRD detection in HPV-independent HNSCC designed specifically for use in the immediate post-surgical window when adjuvant therapy decisions must be made. Accurate MRD identification at this early timepoint has the potential to augment traditional pathology and personalize adjuvant treatment paradigms in HPV-independent HNSCC.

## INTRODUCTION

The incidence of HNSCC continues to increase worldwide and is currently the sixth most common cancer ^1^. While treatment selection considers pathological risk factors such as HPV status, tumor stage or extranodal extension (ENE), 5-year overall survival rates remain low (10-61%, depending on HNSCC subtype and stage) ^2,3^. Treatment approaches such as aggressive adjuvant radiotherapy (RT) or chemotherapy plus radiotherapy (CRT) have been associated with improvements in both PFS and overall survival (OS) in HNSCC patients, but often result in high morbidity, as well as physiological, financial and emotional burden ^4–6^.

There is a critical need to improve treatment selection criteria for patients with HNSCC, improve oncologic outcomes, and limit unnecessary treatment morbidity. While clinicopathologic criteria exist for offering adjuvant radiotherapy or chemoradiation postoperatively for high-risk patients, these criteria are imprecise and have been unchanged for at least 3 decades ^3^. Liquid biopsy, particularly cell-free DNA (cfDNA) based diagnostics in plasma, has been demonstrated to be prognostic, detecting MRD in lung, breast and colorectal cancers ^7–10^. In HNSCC, longitudinal plasma ctDNA detection has been shown to precede the detection of clinical recurrence by surveillance imaging ^11,12^. Clinical use of longitudinal plasma ctDNA is typically employed in a surveillance setting, i.e. to monitor adjuvant treatment response or detect emerging resistance mutations ^13^.

While serial plasma testing achieves high MRD sensitivity in longitudinal analysis (performing correlations with “ever positive” vs. “never positive” over multiple tested timepoints), the landmark (defined as the first post-surgical timepoint only) performance of these tests is not as strong ^14,15^. ctDNA levels may fall below the limit of detection of current technologies in plasma at the landmark timepoint with sensitivities reported at 35-40%^11,16,17^. Additionally, plasma MRD tests frequently initiate testing 1-3 months after surgery ^18,19^, when it is too late to aid in adjuvant treatment decisions ^20,21^. These limitations in plasma-based landmark timepoint testing may be explained by 1) lower ctDNA concentration in the immediate postoperative setting, and 2) the lack of systemic disease in patients undergoing primary surgery.

Given the limitations of plasma-based MRD testing, there has been interest in examining more proximal biofluids, including urine, bronchioalveolar lavage fluid, cerebrospinal fluid (CSF), etc. Tumor proximal biofluids have shown increased sensitivity compared to plasma across multiple cancer types ^22–26^. Our group has pioneered the use of surgical effluent, collected from surgical drains, as a novel proximal biofluid for molecular analysis ^27^. The placement of surgical drains is standard practice following many surgical procedures, including tumor resection, and has been demonstrated to aid in wound-healing and recovery ^28,29^. It is routinely used across cancer types ^30–32^. Although generally discarded, collected surgical effluent has been shown to be reflective of the tissue microenvironment at the surgical site and repair status ^33,34^. It is a mixture of blood, lymphatic exudate, and interstitial fluid, but by 24 hours after surgery, tends to be more lymphatic and interstitial than sanguineous, abbreviated here as “lymph” ^27^. The lymphatic system plays a critical role in the tumor microenvironment and is the primary route of metastatic spread ^35,36^, making it an ideal, but to-date largely unexplored, biospecimen to evaluate early MRD. Previous work from our group demonstrated that HPV viral DNA can be detected in surgical lymphatic fluid at 24-hours post-surgery and that it is prognostic in surgically-treated HPV-associated HNSCC ^27^.

In this study we present a generalizable method for measuring postoperative MRD in HPV-independent HNSCC using lymph ctDNA. Using a tumor-informed, ultra-deep sequencing approach, we demonstrate that lymph ctDNA is more prognostic of recurrence than clinicopathologic features alone and superior to plasma ctDNA from the same timepoint. Our approach overcomes a major limitation of plasma-based MRD assays by measuring proximally to the site of tumor early enough to make adjuvant therapy decisions. The results of this study provide the foundation for interventional clinical trials in HPV-independent HNSCC that use lymph ctDNA in conjunction with pathology to precisely identify and treat patients at higher risk of recurrence through escalation of adjuvant therapy.

## MATERIALS AND METHODS (Fig 1A)

**Figure 1.**
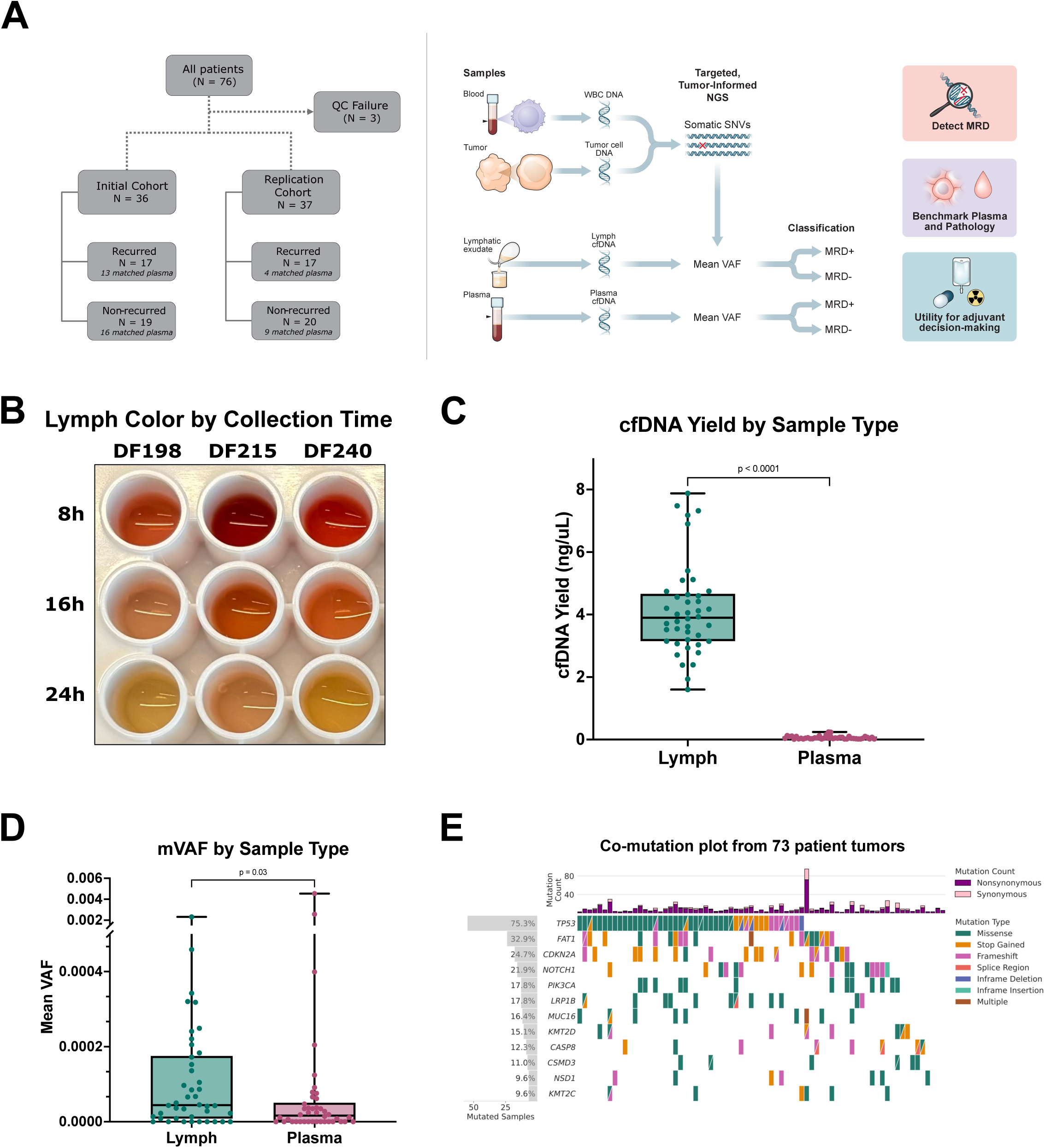
24-hour post-operative lymph is enriched in ctDNA. **A.** Graphical method description. **B.** Representative surgical effluent collected via surgical drains at 8-, 16- and 24-hours post-surgery (n=3 patients). **C.** Higher cfDNA content (ng per μL) of lymph compared to plasma (n=42 patients). **D.** Average variant allele frequency of ctDNA mutations in lymph vs plasma sequenced libraries (n=42 patients). **E.** Co-mutation plot from targeted sequencing of 73 HPV-HNSCC patient tumors (12 most frequently mutated genes shown).

### Ethics Approval and Consent to Participate

The study was conducted in accordance with recognized ethical guidelines put forth in the Declaration of Helsinki. All patients included in this study provided written informed consent to participate. All biospecimens were collected under Washington University School of Medicine Institutional Review Board–approved biospecimen collection protocols #201102323, #201907149, and #201903142 and under the University of Pittsburgh Medical Center Institutional Review Board–approved biospecimen collection protocol UPCI #99-069.

### Study Design and Patient Eligibility

Sixty-two subjects were recruited from Washington University (WashU) between August 2019 and November 2021 and 14 subjects were enrolled from two separate sites within the University of Pittsburgh Medical Center (UPMC) between April 2022 and August 2023. Thirty-six patients were selected at random (blinded to clinical outcomes) from the sequentially enrolled WashU cohort to form the Initial cohort, while 22 WashU patients were held out to be part of the Replication cohort along with the 14 patients from UPMC. Eligibility criteria included: 1) HPV-independent HNSCC, 2) ≥18 years of age, 3) patients undergoing surgical resection of primary tumors with concomitant neck dissection, and surgical drain usage post-operatively. Exclusion criteria included 1) HPV+ HNSCC diagnosis, 2) distant metastatic disease at the time of surgery, 3) synchronous primary tumors, or 4) other primary tumor within the past 5 years. All adjuvant treatment decisions for the patients involved in this study were made without knowledge of assay results. All clinical data were obtained from electronic health records (EHRs) of the enrolled subjects.

Baseline clinical and pathological features were recorded upon patient enrollment in the study. Lymph (n = 76), plasma (n = 42), and peripheral blood samples (n = 76) were prospectively collected at 24 hours post-surgery. Resected tumor samples (n = 76) were also collected. Subsequent follow-up data were collected from the EHRs of the enrolled patients every 4 months and included adjuvant therapy after surgery, treatment outcomes, and dates of disease progression, death, and surveillance follow-up scans.

### Sample collection and Processing

Tumor tissue was collected at surgery, snap-frozen in liquid nitrogen and embedded in optimal cutting temperature (OCT) compound blocks on dry ice. Tissue samples were stored at -80 °C.

All patients underwent Jackson-Pratt (JP) surgical drain insertion during surgery. Lymph was collected from JP drains in 50 mL conical tubes approximately 24 hours after surgery. Collected lymph was transported on ice for processing. Lymph was filtered through a 40 µm cell strainer to remove clumps and debris. K_2_EDTA (8 mM) was added to stabilize cfDNA, inhibit cell coagulation and inactivate nucleases.

K_2_EDTA-stabilized lymph was centrifuged at 2,000 x g for 10 minutes to separate cfDNA-containing supernatant from cells and debris. The clarified supernatant was collected and aliquoted into 2 mL microtubes. Lymph supernatant was stored at –80°C. For bilateral dissections involving two surgical drains, lymph was collected and processed from the side containing biopsy-proven or suspicious lymph nodes.

Peripheral blood was collected in BD Vacutainer K_2_EDTA tubes (BD Biosciences, NJ) concurrently with lymph collection. Collected blood was centrifuged at 1,200 x g for 10 minutes. The plasma (top) layer was transferred to a 15 mL conical tube and centrifuged for 1,800 x g for 5 minutes. Clarified plasma was transferred, mixed and aliquoted into 2 mL microtubes. Plasma was stored at –80 °C. Plasma-depleted whole blood (PDWB) was either mixed and aliquoted into microtubes or buffy coat white blood cells (WBCs) were pelleted. Both PDWB and WBCs were stored at – 80 °C.

### DNA Extraction

Lymph cfDNA was thawed on ice and extracted as follows: 250 µL of lymph was centrifuged at 18,800 x g for 5 minutes at 4 °C and 200 µL transferred to a new tube. The clarified lymph was treated with 3 µL proteinase k (Qiagen, DE) and 10 µL SDS 20%) (Thermo Fisher Scientific, Waltham, MA) and incubated at 60 °C for 20 minutes. Following incubation, cfDNA was isolated from the supernatant after incubation with 0.6x volumetric ratio of SPRISelect beads (Beckman Coulter, Brea, CA). Isolated cfDNA was purified using Zymo Select-a-Size DNA Clean and Concentrator (Zymo Research, Irvine, CA) and eluted in 80 µL of Zymo elution buffer.

Plasma cfDNA was thawed on ice and extracted from 1-4 mL of plasma using the QIAamp Circulating Nucleic Acid Kit (Qiagen, DE). For germline control genomic DNA extraction, 150 µL of PDWB or WBC cell pellet was extracted using the DNeasy blood and tissue kit (Qiagen, DE). Tumor tissue DNA was extracted using the DNeasy blood and tissue kit (Qiagen, DE).

DNA concentrations were quantified using the Qubit HS dsDNA Assay (Thermo Fisher Scientific, Waltham, MA) and an internal lambda DNA control was used to ensure consistent performance (quantified concentration within 20% of known concentration). DNA quality was assessed using the Agilent Tapestation Genomic DNA and cfDNA Analysis kits (Agilent Technologies, Santa Clara, CA). Following DNA extraction, we utilized the following quality-control metrics for inclusion into library prep: cfDNA concentration > 0.4 ng/µL; genomic DNA concentration > 1 ng/µL.

### Library Preparation and Next-Generation Sequencing

Hybrid-capture libraries were prepared using the following method. Genomic DNA was fragmented to 150 bp with the Covaris ME220. 80 ng of cfDNA or 200 ng of genomic DNA was prepared using the xGen cfDNA and FFPE Library preparation kit (Integrated DNA Technologies, Coralville, IA). 600 ng of prepared library was pooled up to 8-plex and hybridized using a standard hybridization reagent kit and a custom hybridization panel covering 699 of the most frequently mutated genes in HPV-independent HNSCC ^37^ (STable 5) (Twist Bioscience, San Francisco, CA). Pre-hybridized and hybridized libraries were assessed using DS1000 DNA Screentape (Agilent Technologies, Santa Clara, CA) and quantified using Qubit HS dsDNA Assay (Thermo Fisher Scientific, Waltham, MA). Coriell DNA samples NA12878 and NA12877 (Coriell Institute, NJ) were used as process controls as follows: NA12877 DNA was spiked into NA12878 DNA at 5% for genomic DNA, and 0.5% for cfDNA, and processed in each library prep batch. The following quality-control metrics for pre-hybridized libraries were used for inclusion into hybridization: Concentration > 10 ng/µL (Qubit HS dsDNA) and average fragment size > 250 bp (Tapestation DS1000), process control > 250 bp average fragment size (Tapestation DS1000), process control concentration >10 ng/µL (Qubit HS dsDNA). The following quality-control metrics of hybridized libraries were used for inclusion into sequencing: Concentration > 10 ng/µL (Qubit HS dsDNA) and average fragment size 300-500 bp (Tapestation DS1000), process control 300-500 bp average fragment size (Tapestation DS1000), process control concentration >10 ng/µL (Qubit HS dsDNA).

Hybridized libraries were sequenced on a NovaSeq 6000 (Illumina, San Diego, CA). The following sequencing run quality-control metrics were used for inclusion into analysis: Q30 > 85%, FASTQ yield > 3Gb (tumor and whole blood) or >180 Gb (lymph or plasma).

### Tumor-informed Variant Calling

Raw reads were demultiplexed using BCL Convert (Illumina, San Diego, CA). Sample matching was performed with a custom pipeline. The pipeline was designed to confirm the identity of FASTQ files by aligning a random subset of reads and comparing highly variable sites with those in BAM files with confirmed identity using Picard (Broad Institute, Cambridge, MA). The comparison generates a LOD score. A score >= 5 indicates a match, and a score < 5 indicates a mismatch or contamination. Consensus read-calling and alignment of reads with UMI family size over 2 were performed as previously described ^27^. Coverage analysis was performed using Picard. Tumor and blood (PDWB or WBC) samples with median target coverage lower than 250x were excluded from further analysis. Lymph and plasma samples with median target coverage lower than 2500x were excluded from further analysis.

Tumor variant calling was performed as previously published ^27^. A panel of normals was created with 50 whole blood samples following GATK best practices. From these variants, somatic mutations on coding sequences were identified using Ensembl Variant Effect Predictor annotation. Somatic mutations with depth <50x or variant allele fraction (VAF) <5% were excluded. Somatic mutations on sex chromosomes were also excluded. Matched whole blood samples were used to filter out germline mutations or mutations driven by clonal hematopoiesis. All remaining somatic mutations identified in tumor were directly genotyped in matched lymph and plasma samples. Mutated gene frequencies were estimated in 415 HPV-independent HNSCC tumor samples from The Cancer Genome Atlas (TCGA) and compared with mutational landscape of the tumor and lymph samples in this study.

A base-specific error model (BEM) was estimated at each somatic mutation position to quantify the background noise for single nucleotide variants (SNVs). Models for lymph were built using a total of 91 high-quality lymph samples sequenced to a median coverage of 6,328x (range 2,544x to 11,580x), while models for plasma were built using a total of 49 plasma samples sequenced to median coverage of 6,191x (range 2,911x to 13,675x). BEM was fit by Weibull distribution if more than 5 non-zero non-reference alleles were observed at a tumor variant position across the reference samples ^38^. Otherwise, Gaussian distribution was used to fit the BEM. All models were fit using the SciPy package.

For each model, the BEM cutoff was defined as the 75th percentile of the distribution, which was adjusted accounting for zero-inflated reference data. We used 75th percentile to control the false discovery rate (FDR) so that true variants would not be filtered out based on our observations through reference data. For indels, the cutoff was defined as 0.1% considering the minimal coverage required to achieve at least 2 non-reference reads. For each variant in lymph and plasma samples, VAF was set to 0 if not greater than the BEM cutoff for SNVs and indel cutoff for indels. Mean VAF was calculated for each lymph and plasma sample using all somatic mutations that passed filtering in the tumor of the patient. Somatic variants that were not detected in fluid were input into the mean VAF calculations as “0”. For the contrived process control, externally validated high confidence SNVs in NA12877 ^39^ were quantified. The metric used was ≤20% of detected VAF variation from the expected VAF, based on NA12877 DNA spike-in percentage.

### Calibrated MRD cut-off

Due to differences in observed tumor mutation count and sequencing coverage, a unified MRD cutoff would bias the MRD prediction for patients with higher mutation rates or lower coverage. Therefore, we applied correction factors to the cutoff (C) to account for tumor mutation count (N_t_) and mean coverage depth (D_m_) for each lymph or plasma sample using the simple formula below:

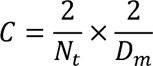

cfDNA samples with mean VAF greater than the cutoff were considered MRD positive, and those with mean VAF less than or equal to the cutoff were considered MRD negative.

To demonstrate the robustness of the calibrated MRD cutoff, we randomly downsampled one lymph sample with high observed tumor mutation count and coverage (SFIG 1A-C). Three technical replicates were created for downsampling to demonstrate consistency with different downsampling seeding. These data indicate that with the use of the coverage- and mutation count-corrected cutoff, we were able to make consistent, accurate MRD calls despite variation in the observed tumor mutation count and achieved sequencing coverage of the lymph sample.

### Logistic Regression Model of Pathologic Features

A logistic regression model including 5 high-risk pathologic features (margins, extranodal extension, perineural invasion, lymphovascular invasion, and regional node involvement) was built as the baseline. All 5 pathologic features were binary. Another logistic regression model combining lymph ctDNA status with these 5 pathologic features was built for comparison. To avoid overfitting, 5-fold cross validation was repeated 100 times to calculate the average prediction scores for each patient. Youden’s index was calculated to serve as the MRD cutoff for prediction scores. For each model, survival analysis was performed based on average prediction scores to examine the model performance.

### Detection Rate of Mutations at Ultra-low VAF

DNA from two Coriell cell lines (NA12877 and NA12878) was prepared with the custom hybrid-capture panel, sequenced and aligned to the human reference genome, using the methodology described above for tissue and lymph samples. NA12877 was downsampled to 1/1000, 1/10000, and 1/100000 reads first, and then merged with NA12878 to create 3 samples at different mixing ratios. Three replicates were created at each downsampling ratio, resulting in a total of 9 samples. Variant calling was performed on known heterozygous and homozygous mutations of NA12877 in these 9 samples to determine the sensitivity of the assay at ultra-low VAFs.

### Statistical Analysis

Following binary prediction by the models as described above, Kaplan-Meier estimator with log-rank test and Cox proportional-hazards regression models were used for survival analysis. For clinical features, missing values were imputed by mode ^40^ except for tumor size where median was used in covariate analysis (STable 4) and Cox regression analysis (STable 8-9). Not all high-risk features had missing values. Sensitivity and specificity were calculated based on ultimate disease progression status. Wilcoxon signed-rank test was used for group comparisons. Student’s t-test, Fisher’s exact test, and Spearman’s correlation were also utilized as appropriate. A p-value ≤ 0.05 was considered statistically significant for all statistical tests. Python 3.8.10 was used for all statistical analyses.

### Orthogonal Validation

Labcorp Plasma Detect is a clinically validated, tumor-informed MRD test using ctDNA whole-genome sequencing (Baltimore, MD), and was utilized for orthogonal validation ^41^. Briefly, whole-genome sequencing was performed from tumor tissue, PDWB, plasma, and lymph-derived DNA using up to 200 ng, 50 ng, 10 ng, and 10 ng of input material, respectively, using the Illumina NovaSeq 6000 platform. Tumor-specific SNVs were identified from tumor and germline datasets, from which a candidate variant set was used to determine the presence of ctDNA within lymph or plasma DNA through a random forest machine learning model. Specifically, candidate tumor-specific SNVs identified in the test sample were scored (ranging from 0 to 1) using a random forest machine learning algorithm trained using the caret package (v6.0.90) within the R statistical computing environment (v4.1.1). Model training occurred independently of the clinical cohort, using a combination of contrived reference cell line and noncancerous donor specimens, ultimately leveraging 500 trees, approximately 7-8 levels, and approximately 150,000 leaves ^41^. To avoid overfitting, model training utilized 5-fold cross-validation and limited the number of selected variables per split procedure (hyperparameter mtry) to the square-root of the total number of input features. Finally, variants present in properly paired mapped fragments with a random forest score >0.25 were further assessed, requiring an alternate read-mapping quality ≥30 and a read-based mutation rate ≤5. ctDNA status was determined based on the aggregate level of signal observed across all candidate variants compared to a commercially procured reference population of noncancerous donor plasma samples [n = 80, obtained under Institutional Review Board approval from Discovery Life Sciences (Alabama, USA)] and a cutoff of one standard deviation above the maximum observed value was required to report a plasma or lymph sample as ctDNA positive. An estimated tumor fraction was then calculated for each positive sample based on the aggregate variant allele observations observed as a proportion of the total unique coverage of all individual tumor-specific variants assessed.

Lymph, plasma, and blood (PDWB or WBC) were collected 24 hours postoperatively from 38 HPV-independent HNSCC patients along with resected tumor specimens. Patient-matched tumor, germline (blood), and cfDNA from lymph or plasma samples were evaluated for orthogonal validation and sequenced at approximately 150x, 30x, and 30x depth, respectively. 5 tumor and 1 lymph samples failed whole genome library preparation quality-control (4.6 nM and 1.4 nM, respectively); 2 lymph samples completed Plasma Detect processing, but did not complete hybrid-capture processing due to insufficient DNA. Therefore, 30 patients remained, of which, 13 patients experienced disease recurrence and 17 demonstrated no evidence of disease with >1 year of follow-up. Twenty-four of these patients with lymph samples processed by Labcorp Plasma Detect also had matched plasma samples available to enable direct lymph vs. plasma comparisons. KM estimator with log-rank test and Cox proportional-hazards models were used for survival analyses.

## RESULTS

### Study Population

76 HPV-independent HNSCC patients were initially included in the study (Fig 1A). One patient was excluded due to a blood sample mismatch and two patients were excluded because tumor samples failed sequencing, resulting in a total study population of 73 patients. Of the 73 patients, 36 patients were studied in an initial cohort and 37 were reserved for the replication cohort. Extranodal extension (ENE), immunotherapy, and smoking packs per year had modest differences between the two groups; there were no other clinical or demographic features with significant differences between the initial and replication cohorts (STable 1-4). The median age of the study population was 63. Forty-nine (67.1%) were male and 52 (71.2%) patients received adjuvant therapy (1.4% chemotherapy, 46.6% radiotherapy, 20.5% chemoradiation, 2.7% immunotherapy). The median follow up was 37.4 months. Thirty-four out of 73 patients experienced disease recurrence (REC) (17 initial cohort and 17 replication cohort) and 39 out of 73 patients had no evidence of disease (NED) (19 initial and 20 replication cohort) (Fig 1A). Forty-two patients had matched plasma samples available (29 initial cohort,13 replication cohort).

### Lymph and Plasma sample composition at 24 hours post-surgery

Surgical drain fluid composition changes over time, transitioning from sanguinous to more serous (lymphatic and interstitial) fluid at 24 hours (Fig 1B). Indeed, we observed distinct cfDNA characteristics in postoperative lymph. cfDNA in lymph was 65 times more concentrated than plasma (mean 4.14 ng per μL of lymph, mean 0.06 ng per μL plasma, *P* < 0.0001) (Fig 1C). Additionally, lymph cfDNA displays a distinct nucleosome distribution and a higher dinucleosome fraction compared to plasma ^27^. The mean ctDNA allelic fraction was 2 times higher in lymph than in plasma samples (lymph = 0.004 ± 0.036%; plasma = 0.002 ± 0.078%; *P* = 0.03, n = 42) (FIG 1D). Taken together, lymph represents a differentiated biofluid with substantially higher ctDNA content than plasma.

### Lymph but not Plasma ctDNA Correlates with HNSCC Recurrence at the Landmark Timepoint

To maximize sensitivity and specificity while still achieving a <2 week result turnaround time, we employed an approach in which all analytes are sequenced using the same panel, but the variant-calling is performed in a tumor-informed manner. Tumor, blood, lymph, and plasma samples were each sequenced using a custom 699-gene panel comprised of the most frequently mutated genes in HPV-independent HNSCC (STable 5) ^37^. This ultra-deep-coverage next-generation sequencing assay is demonstrated to routinely detect variants down to 1:100,000 (SFIG 1D). Tumor and whole blood libraries were sequenced to a median of 533x deduplicated coverage (range 249x - 2709x). Lymph and plasma cfDNA libraries were sequenced to a median of 6,322x deduplicated coverage (range 2,544x - 13,675x). Variant-calling was then performed in a tumor-informed fashion, where patient-specific somatic variants (median 9, range 2 - 95) detected in tumor were directly genotyped to examine those positions in lymph and plasma. Lymph samples were classified as positive if the mean ctDNA VAF exceeded the prespecified cut-off after applying a non-linear correction for coverage and mutation load variation (see Methods). Key metrics for all lymph samples are summarized in STable 6. We evaluated the mutational landscape of tumor (STable 7, SFIG 2) and lymph (SFIG 3) samples, finding that mutated gene frequencies were comparable to TCGA data of HPV-independent HNSCC (FIG 1E and SFIG 2).

We performed survival analysis using the determined MRD status from both lymph and plasma samples from the initial study cohort. We found that lymph ctDNA could accurately identify HNSCC recurrence (n = 36, sensitivity = 76%, specificity = 63%; *P* = 0.01, Log-rank test for all subsequent *P* unless specified otherwise) (FIG 2A). The hazard ratio (HR) for disease recurrence in lymph ctDNA+ patients was 3.8, indicating that disease recurrence was nearly 4 times more likely among patients with detectable lymph ctDNA than among those without during the follow-up period. We next attempted to replicate these results in an independent cohort of 37 patients. Again, lymph ctDNA positivity accurately identified HNSCC recurrence (sensitivity = 65%, specificity = 70%; *P* = 0.04, HR = 2.8) (FIG 2B), suggesting that lymph profiling is a reproducible approach to detect MRD 24 hours after surgery.

**Figure 2.**
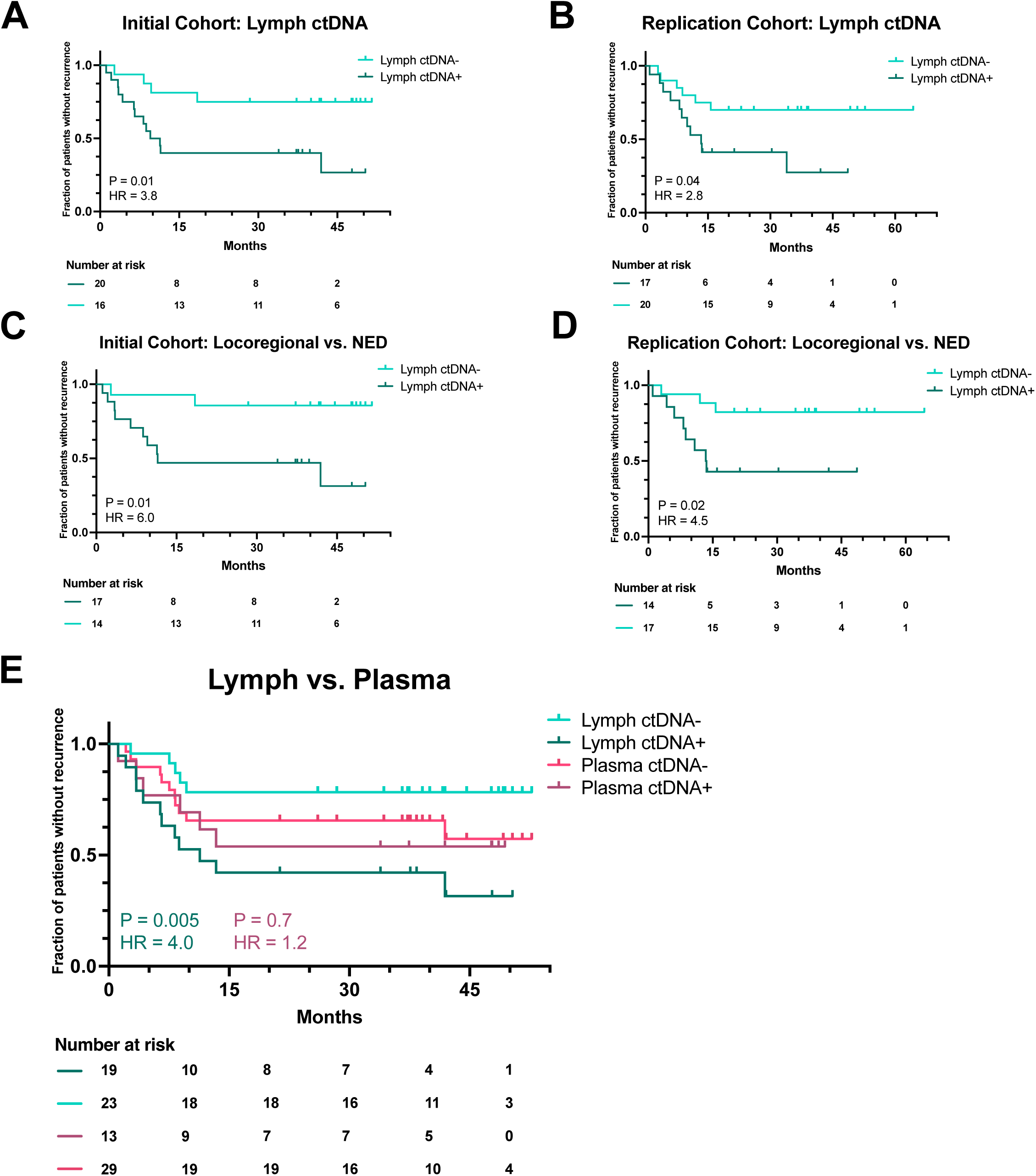
Lymph but not plasma ctDNA correlates with HNSCC recurrence. **A.** KM PFS Analysis by Lymph ctDNA status for Initial Cohort (n=36), sensitivity = 76%, specificity = 63%; Logrank *P* = 0.01, HR = 3.8 (95% CI, 1.2-11.8) . **B.** KM PFS Analysis by lymph ctDNA Status for Replication Cohort (n=37), sensitivity = 65%, specificity = 70%; Logrank *P* = 0.04, HR = 2.8 (95% CI, 1.0-7.7). **C.** KM PFS Analysis by lymph ctDNA status for locoregional recurred and NED patients in Initial Cohort (n=31), sensitivity = 83%, specificity = 63%; Logrank *P* = 0.01, HR = 6.0 (95% CI, 1.3-27.7). **D.** KM PFS Analysis by lymph ctDNA status for locoregional recurred and NED patients in Replication Cohort (n=31), sensitivity = 73%, specificity = 70%; Logrank *P* = 0.02, HR = 4.5 (95% CI, 1.2-17.1). **E.** KM PFS analysis by lymph ctDNA status vs. plasma in matched patient cohort (n=42), lymph sensitivity = 71%, specificity = 72%; Logrank *P* = 0.005, HR = 4.0 (95% CI, 1.4-11.4); plasma sensitivity = 35%, specificity = 72%; Logrank P = 0.70, HR = 1.2 (95% CI, 0.5-3.3).

To evaluate the potential benefit of a proximal fluid on local relapse patterns, we next compared patients with locoregional relapse and NED patients in both the initial and replication cohorts. Detection of ctDNA in lymph was strongly associated with recurrence in both cohorts (Cohort 1: n = 31, sensitivity = 83%, specificity = 63%; *P* = 0.01, HR = 6.0; Cohort 2: n = 31, sensitivity = 73%, specificity = 70%; *P* = 0.02, HR = 4.5) (FIG 2C-D). The reproducible enhanced sensitivity for locoregional recurrence observed in these cohorts is consistent with the hypothesis that a proximal fluid will be enriched for tumor-associated analytes in the setting of local tumor growth.

Forty-two of the patients across these cohorts had matched plasma drawn 24 hours after surgery. Applying the same ultra-sensitive NGS approach used for lymph, plasma was not predictive of MRD at this timepoint (sensitivity = 35%, specificity = 72%; *P* = 0.7, HR = 1.2; positive predictive value (PPV) = 46%, negative predictive value (NPV) = 62%) (Fig 2E). Lymph correctly identified MRD twice as often in the matched cohort (sensitivity = 71%, specificity = 72%; p = 0.005, HR = 4.0; PPV = 63%, NPV = 78%). Plasma ctDNA performed worse in prognostication of locoregional disease (n = 34, sensitivity = 33%, specificity = 72%; *P* = 0.86, HR = 1.1) (SFIG 4A), in line with ctDNA work involving other proximal biofluids ^23,24^. Reflective of overall concordant biology, the Spearman’s rank correlation coefficient between VAFs in the matched cohort showed a weak correlation for lymph and plasma samples (*P* = 0.0005, r^2^ = 0.03) (SFIG 4B).

### Lymph ctDNA independently outperforms plasma through orthogonal validation

To complement these findings, we sought to orthogonally validate these results through independent analysis of Cohort 1 (plus two additional patients) with the clinically-validated Labcorp Plasma Detect (PD) MRD assay, which represents a bespoke, tumor-informed, whole-genome sequencing approach. The Spearman’s rank correlation coefficient between lymph mVAF and PD ctDNA level and Fisher’s exact test between MRD prediction by lymph mVAF and PD ctDNA showed excellent concordance for lymph samples evaluated through both methods (mVAF vs ctDNA level r^2^ = 0.49, *P* < 0.0001; Fisher’s exact test *P* = 0.0001) (SFIG 4C-D).

Lymph-derived ctDNA status determined by PD again accurately identified disease recurrence (n = 30, sensitivity = 50%, specificity = 88%; *P* = 0.003, HR = 4.8), consistent with our previous results using targeted sequencing applied to the same patient cohort (sensitivity = 71%, specificity = 75%; *P* = 0.006, HR = 4.5) (SFIG 4E-F). In the sub-cohort of 14 N0 stage patients, the PD assay applied to lymph cfDNA also demonstrated strong association with recurrence (sensitivity = 75%, specificity = 90%; *P* = 0.004, HR = 14.5) (SFIG 4G).

We then performed survival analyses using the PD ctDNA MRD status derived from plasma samples for 24 patients where matched lymph samples were available. Lymph MRD status again accurately identified disease recurrence in this subgroup (n = 24, sensitivity = 45%, specificity = 92%; *P* = 0.003, HR = 6.0) (SFIG 4H) and also outperformed plasma ctDNA MRD status at this timepoint (n = 24, sensitivity = 27%, specificity = 92%, *P* = 0.03, HR = 4.2) (SFIG 4I), replicating the observed reduced performance of plasma for determination of MRD status at the 24-hour post-surgery timepoint.

### Lymph ctDNA outperforms individual pathology features and shows synergy in a combination model

We compared relevant clinical features (e.g. ENE status, tumor stage, etc.) between our two cohorts and demonstrated minimal difference between these populations (STable 4). Having established their similarity, we then pooled the cohorts into a single 73-patient group to better power subgroup analyses. In aggregate, lymph ctDNA in the pooled cohort demonstrated 71% sensitivity and 67% specificity to detect HNSCC recurrence (*P* = 0.001, HR = 3.2) and again demonstrated enhanced locoregional performance (n = 62, sensitivity = 78%, specificity = 67%, *P* = 0.0004, HR = 5.1) (FIG 3A, SFIG 5A). Multivariate Cox regression analysis was performed with lymph ctDNA status and relevant clinical features for PFS (STable 8). Only lymph ctDNA MRD detection and node-positive status were associated with significantly worse PFS, indicating minimal confounding effects for lymph ctDNA due to clinical features. Similar findings were observed for OS, with lymph ctDNA MRD+ patients experiencing significantly worse survival, even when accounting for clinical covariates (SFIG 5B and STable 9). Enhanced locoregional performance (sensitivity = 79%, specificity = 58%, *P* = 0.014, HR = 4.4) was again observed for OS (SFIG 5C). We evaluated the prognostic value of lymph ctDNA in lower-risk pathologies as well, including patients without ENE, N0 patients, and patients with T1-T2 disease (FIG 3B, SFIG 5D-E). Lymph ctDNA performed consistently in the N0 (*P* = 0.11, HR = 3.5) and T1-T2 (*P* = 0.3, HR = 1.9) population, similar to the full cohort (albeit not significant given this small N), suggesting lymph ctDNA detection may be a relevant biomarker in earlier stage patients (SFIG 5D-E). Furthermore, lymph ctDNA was strongly associated with recurrence in patients without ENE (n = 50, sensitivity = 80%, specificity = 67%; *P* = 0.001, HR = 5.2) (FIG 3B), demonstrating that lymph ctDNA can identify patients at higher risk of recurrence despite their having lower risk pathology.

**Figure 3.**
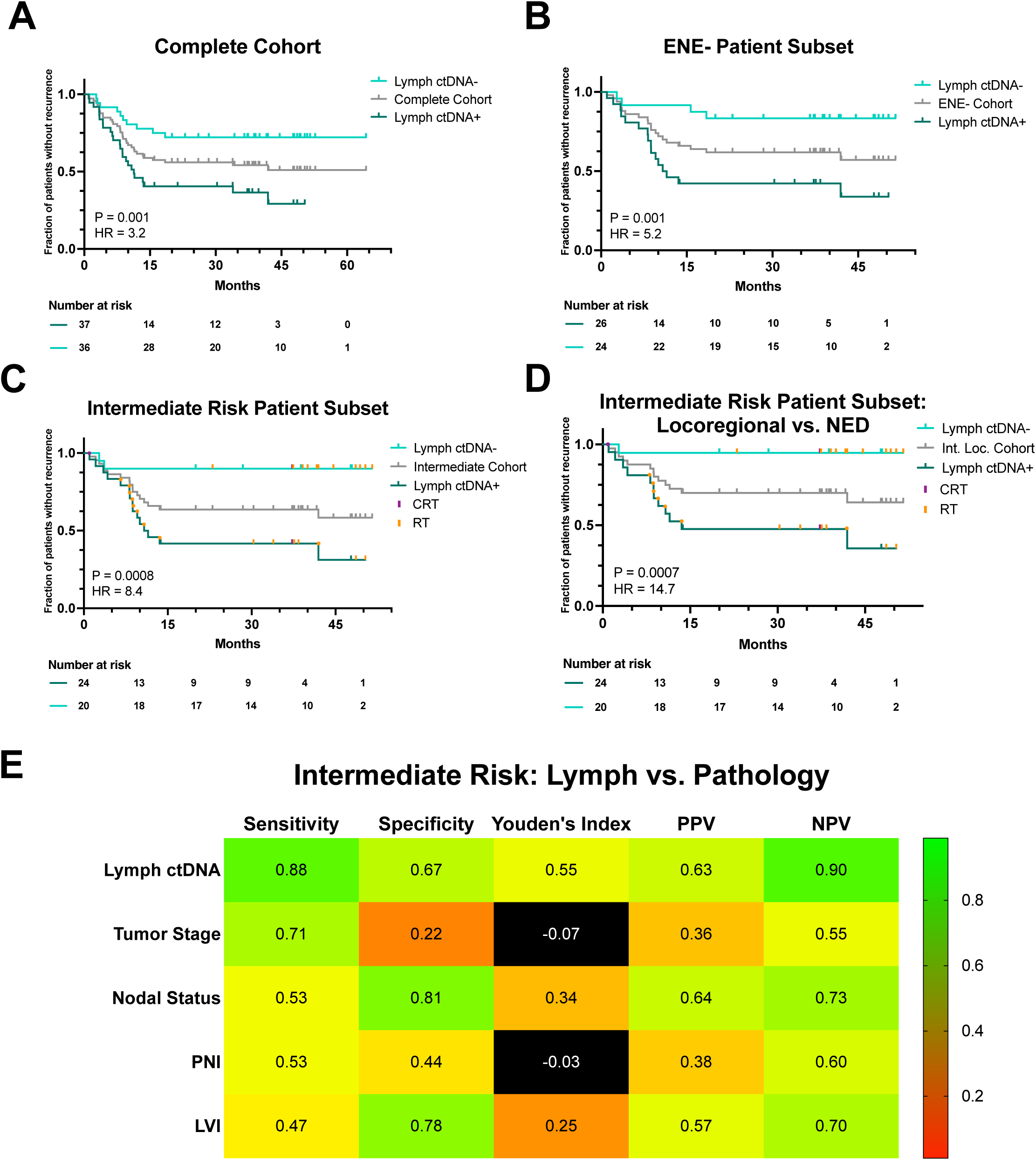
Lymph ctDNA further stratifies HNSCC recurrence in intermediate-risk patients (ENE and positive margins negative with at least one other adverse pathology feature: >T2, >N1, PNI, or LVI). **A.** KM PFS analysis by lymph ctDNA status for Complete Cohort (n=73), sensitivity = 71%, specificity = 67%, Logrank *P* = 0.001, HR = 3.2 (95% CI, 1.5-6.8). **B.** KM PFS analysis by lymph ctDNA status for ENE-negative patients (n=50), sensitivity = 80%, specificity = 67%, Logrank *P* = 0.001, HR = 5.2 (95% CI, 1.7-15.7). **C.** KM PFS analysis by lymph ctDNA status for intermediate-risk patients (n=44), sensitivity = 88%, specificity = 67%, Logrank P = 0.0008, HR = 8.4 (95% CI, 1.9-37.1). **D.** KM PFS analysis by lymph ctDNA status for locoregional recurred and NED patients with intermediate risk (n=40), sensitivity = 92%, specificity = 67%, Logrank *P* = 0.0007, HR = 14.7 (95% CI, 1.9-113.4). **E.** Accuracy of lymph ctDNA compared to pathology biomarkers for intermediate-risk patients.

We next sought to benchmark the prognostic value of lymph ctDNA compared to individual adverse pathologic features that are considered during treatment selection [tumor stage, TNM stage, final margins, ENE, regional node involvement, perineural invasion (PNI), and lymphovascular invasion (LVI)]. Final margins, regional nodal status, and LVI were statistically associated with disease recurrence in this cohort, but with the exception of residual cancer left behind (positive tumor margins), none were more prognostic of recurrence than lymph ctDNA (SFIG 5F-L). We next performed a simple logistic regression model combining five key pathologic factors (Methods). This model was significantly associated with recurrence (sensitivity = 44%, specificity = 85%; *P* = 0.002, HR = 2.8) but underperformed compared to lymph ctDNA alone (SFIG 5M).

A differentiating feature of a lymph-based MRD approach is that results could be available shortly after surgery and in the same timeframe as surgical pathology. Therefore, we sought to determine how lymph ctDNA interacts with intermediate-risk pathology feature classification by using a logistic regression model, incorporating the 5 pathologic features examined above plus lymph ctDNA. When lymph ctDNA was considered alongside these pathology features, recurrence detection was improved compared to pathology or lymph ctDNA status alone (sensitivity = 68%, specificity = 74%; *P* = 0.0003, HR = 3.5), suggesting potential synergy between pathologic and molecular features (SFIG 5M).

### Potential Predictive Applications of Lymph ctDNA in Intermediate-Risk HPV-independent HNSCC

Under NCCN guidelines ^42^, patients with the highest risk features (ENE or positive margins) are recommended to receive maximal adjuvant therapy after surgery (chemoradiation; CRT), while those without ENE or positive margins but who do have at least one other adverse pathologic feature (>T2, >N1, PNI, or LVI) are recommended to receive radiotherapy (RT) and to “consider systemic therapy”.

Given the synergy we observed between pathology and lymph ctDNA, we tested whether lymph ctDNA has utility to further stratify recurrence risk in patients with intermediate-risk pathology. In this sub-cohort, we observed that patients who are positive for lymph ctDNA at 24 hours are significantly more likely to have disease recurrence (n = 44, sensitivity = 88%, specificity = 67%; *P* = 0.0008, HR = 8.4) (FIG 3C). Additionally, when we looked at patients who received RT alone and later recurred, we saw that 100% (9/9) were positive for lymph ctDNA at 24 hours (FIG 3C). We again observed enhanced locoregional performance (sensitivity = 92%, specificity = 67%, *P* = 0.0007, HR = 14.7) in this sub-cohort (FIG 3D). Indeed, lymph ctDNA yielded a higher accuracy than any of the pathology biomarkers that were available at this early timepoint (Youden’s index = 0.55 vs. -0.07 – 0.34) (FIG 3E). This suggests the potential for using a molecular test to further stratify intermediate-risk patients who might benefit from treatment intensification after surgery. These findings provide rationale for future interventional studies incorporating lymph ctDNA status into adjuvant treatment decision-making for HPV(-) HNSCC patients undergoing surgery.

To illustrate the potential utility of lymph ctDNA testing to impact clinical care, we present two case studies that focus on patients with pathologically intermediate-risk disease who underwent curative-intent tumor resection surgery. Patient DF067 (FIG 4A) was T2N0M0 with LVI and received 70 Gy of adjuvant RT but no chemotherapy. One day after surgery, postoperative lymph was positive for ctDNA. Thirteen months later, DF067 suffered a locoregional recurrence in the right palate.

**Figure 4.**
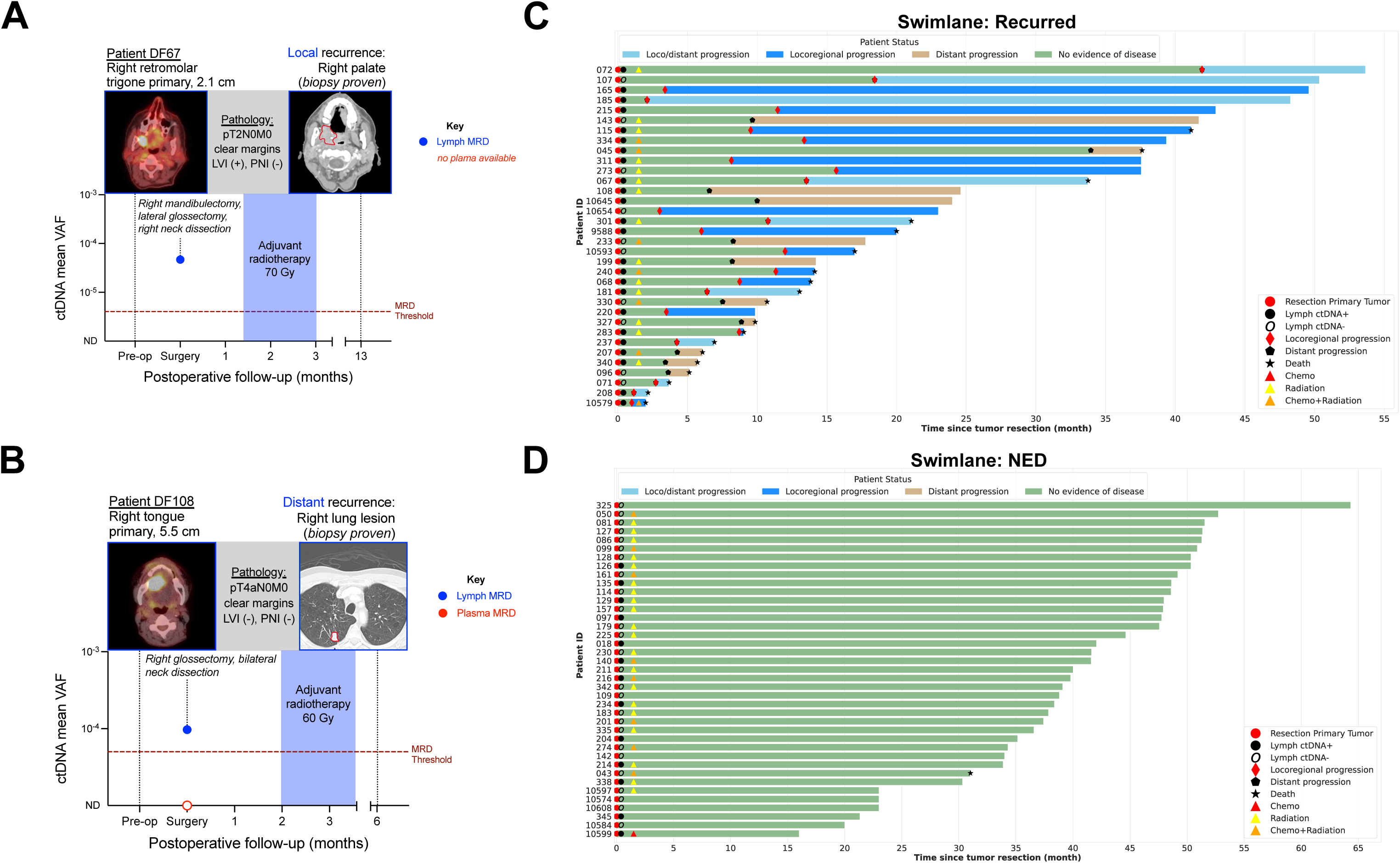
Lymph ctDNA can inform adjuvant treatment decision-making. **A.** Clinical vignette of patient DF67 with positive lymph ctDNA. Patient received 70 Gy of adjuvant RT but no chemotherapy. Patient experienced locoregional recurrence in the right palate 13 months later. **B.** Clinical vignette of intermediate-risk patient DF108 with T4N0M0 oral HNSCC, who was managed with radiotherapy. Lymph ctDNA was positive but plasma ctDNA was not. Patient experienced distant recurrence (lung) 6 months after surgery. Representative lung CT image shown as previously published ^49^. **C.** Swimmer plot of recurred patients by patient ID (n=31) describing lymph ctDNA status, treatment course, recurrence type and outcome. **D.** Swimmer plot of non-recurred patients by patient ID (n=42) describing lymph ctDNA status, treatment course, recurrence type and outcome.

Patient DF108 (FIG 4B) was T4N0M0 with no evidence of other high risk pathological features and received 60 Gy adjuvant RT but no chemotherapy. The 24- hour lymph sample was positive for ctDNA while the 24-hour plasma sample was negative. Six months after surgery, the patient recurred distantly in the right lung. In both cases, postoperative lymph ctDNA tests were consistent with eventual relapse. These data illustrate that some patients with low/intermediate-risk pathology will still experience relapses that pathology alone cannot predict but are detectable at the molecular level in lymph fluid. Lymph ctDNA represents a novel and complementary facet to relapse risk prediction that in the future could be used as an adjunct to pathology to better tailor adjuvant therapy selection.

## DISCUSSION

Our work is the first to establish the potential utility of surgical lymphatic fluid as a proximal bioanalyte for measuring MRD for use in adjuvant therapy decision-making in HPV-independent HNSCC. In this multi-institutional cohort of patients, we show that those positive for lymph ctDNA at the landmark postoperative timepoint were significantly more likely to experience recurrence. The results were most pronounced for intermediate-risk HPV-independent HNSCC, where the need for clarity and precision in adjuvant therapy decision-making is highest. We demonstrate that lymph has a 65-fold higher cfDNA yield per mL and a 2x higher tumor allelic fraction than time-matched plasma. With a 130-fold advantage in available tumor-derived DNA, lymph ctDNA tests could disproportionately benefit from emerging ultra-sensitive NGS methods with less than 1 part per million detection ^43,44^ and beyond. Indeed, the fundamental properties of lymph have the potential to overcome the sensitivity challenges faced by plasma testing in the immediate postoperative setting without changes to standard clinical workflows or additional invasive procedures. As an NGS-based MRD test, the cost and resources needed for lymph testing is similar to current plasma-based tests on the market.

In our initial cohort, we found that lymph ctDNA was significantly associated with recurrence detection, findings that then replicated in an independent multi-site cohort. Strikingly, lymph ctDNA accurately identified HNSCC recurrence with a sensitivity of 71% and a specificity of 67% (*P* = 0.001) in the full cohort (FIG 3A), whereas plasma was not significant at this timepoint with a sensitivity of only 35%, identifying MRD in less than half as many patients.

Plasma-based MRD assays have historically struggled to detect locoregional recurrence ^45^. Our results suggest that lymph effectively overcomes this difficulty for HPV(-) head and neck cancer. Local recurrence within 1 year was 5 times more likely among patients with ctDNA-positive lymph samples than among those with ctDNA-negative lymph samples. Accurate MRD identification at an immediate postoperative timepoint in patients with locoregional disease suggests that postoperative lymph analysis may have the potential to detect residual cancer before it spreads beyond the original tumor site and adjacent lymph nodes.

We also observed that lymph outperforms most individual pathology features or a combination of them at identifying recurrence. Lymph ctDNA correlated with recurrence in patients with intermediate-risk pathology with a sensitivity of 88% and a specificity of 67% (*P* = 0.0008). This suggests that a molecular biofluid test has potential to further stratify recurrence risk beyond traditional pathology. Given the 24-hour sampling, lymph ctDNA test results could be reported <2 weeks after surgery, on a similar time scale as traditional pathology reports. This combination of complementary information and synchronous timing indicates that postoperative lymph testing could enable MRD findings to be used in concert with traditional pathology measures to provide more accurate, personalized adjuvant treatment decisions and be readily incorporated into multidisciplinary tumor board discussions.

The results of this study highlight the potential value of lymph ctDNA testing over plasma in the early postoperative period, but the temporary nature of surgical drains limits longitudinal surveillance using this approach alone. MRD detection using lymph and plasma could be complementary, however: for example, an initial lymph test 24 hours after surgery to enable sensitive MRD results in time to inform adjuvant therapy decisions could be combined with longitudinal plasma (or other renewable biofluid) testing to monitor ongoing patient response to therapy and identify recurrence at later timepoints.

An important limitation of the current work is that as an observational study, no patients were treated based on their lymph ctDNA results. The majority of patients (73.3%) received adjuvant therapy according to the standard of care, regardless of their lymph ctDNA status. As a result, we are unable to discriminate patients who were misclassified as ctDNA+ (technical false positives) from those who were biologically ctDNA+ at 24 hours but responded to adjuvant therapy and therefore did not relapse. Therefore, specificity is likely underestimated from both lymph and plasma given that samples were collected prior to adjuvant therapy administration. Indeed, 9 out of 13 misclassified lymph ctDNA+ NED patients received adjuvant therapy. Reported RT and CRT response rates ^46^ would suggest that some of these apparent “false positives” at 24 hours were in fact patients who responded to subsequent adjuvant treatment and therefore had favorable outcomes despite having MRD after surgery. We also observed 4 false positives among patients who received no reported adjuvant therapy. There are several possible explanations for this, including: limited follow-up time (3 out of 4 had <3 years of follow-up), possible inaccuracy or incompleteness of treatment records and/or recurrence data for patients receiving follow-up care outside of the collaborating tertiary-care academic medical center, and the choice of MRD cutoff (which was tuned to maximize sensitivity). These hypotheses can be tested in future prospective clinical trials. As a biofluid, the limited availability of surgical drain fluid (given its novelty and lack of inclusion in general banking protocols) restricted the sample size in this study, particularly for subset analyses. Future studies with larger cohort sizes will enable validation of current findings and further analyses into relevant patient subsets.

In this cohort, we observed that lymph ctDNA is prognostic of PFS in intermediate-risk HPV-independent head and neck cancer patients who did not receive maximum adjuvant therapy (CRT). This, combined with accurate MRD detection in patients with lower-risk pathologic features, provides proof-of-principle that postoperative lymph MRD testing has the potential to better stratify patients to receive more personalized adjuvant treatment. Clinical trials have demonstrated that escalating treatment from RT to CRT can improve median OS by 3.3 years ^3^, but the addition of chemotherapy comes with significant treatment morbidities ^47^. A treatment stratification paradigm incorporating lymph ctDNA testing could help improve outcomes and limit overtreatment by targeting therapy to those most at risk of relapse. The next step to demonstrate the utility of a lymph ctDNA-centric approach is validation in interventional clinical trials. This may include, for instance, patients with intermediate pathologic risk who are stratified to escalated adjuvant therapy based on a positive lymph ctDNA result.

Indeed, lymph ctDNA detection may address a critical yet unresolved need in HPV-independent HNSCC treatment. The unique sensitivity at 24-hours post-surgery may confer the added benefit of aligning to total package time for adjuvant treatment decision-making, which is crucial for maximizing patient survival^48^, but remains a limitation of plasma-based ctDNA MRD testing. Future work will focus on investigating these aspects in prospective studies.

In conclusion, we introduce postoperative lymph ctDNA as a novel, proximal, and early source to detect MRD with the potential to introduce more precision into adjuvant therapy decision-making for HPV-independent HNSCC, especially for intermediate risk patients. Advances in liquid biopsy technology may allow a biofluid that has been overlooked for decades to enable precision adjuvant therapy decision-making and achieve the best oncologic outcomes possible.

## Data Availability

All data produced in the present study are available upon reasonable request to the authors

## ACKNOWLEDGEMENTS

The authors thank Katie Campbell for helpful feedback on the manuscript and Veronica Falconieri Hays for illustration assistance on Figure 1A.

---

**Supplementary Figure 1.**
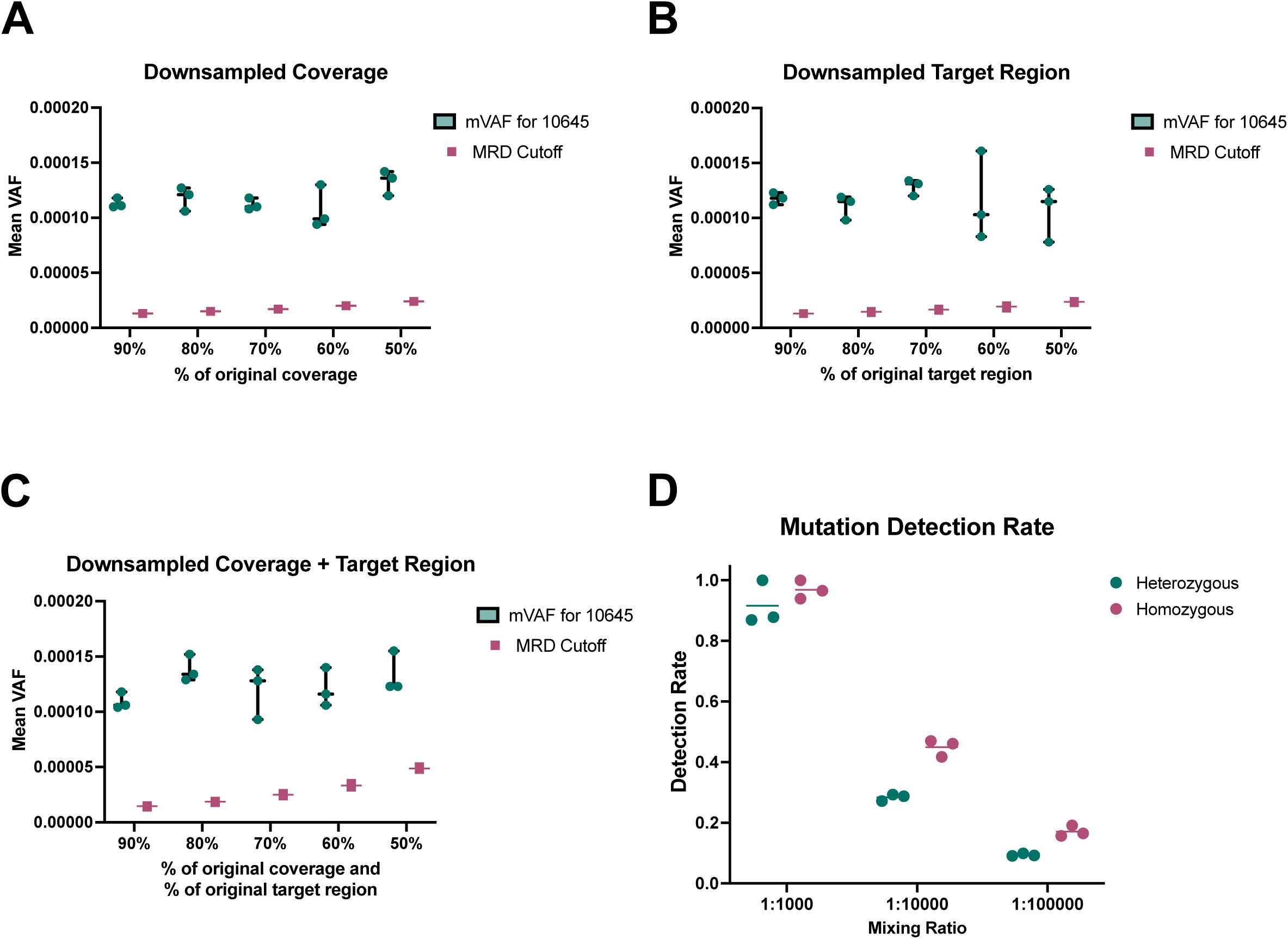
Mutation Detection in Methods. **A.** Comparison of calibrated cut off and mVAF for patient 10645 at various coverage levels (90-50%). **B.** Comparison of calibrated cut off and mean VAF for patient 10645 at various target region sizes (90-50%). **C.** Comparison of calibrated cut off and mVAF at various coverage and target region sizes (90-50%) for patient 10645. **D.** Mutation detection rate by different mixing ratios, for known homozygous mutations, 97% mutations at 0.1% VAF, 45% mutations at 0.01% VAF, and 17% mutations at 0.001% VAF were detected respectively.

**Supplementary Figure 2.**
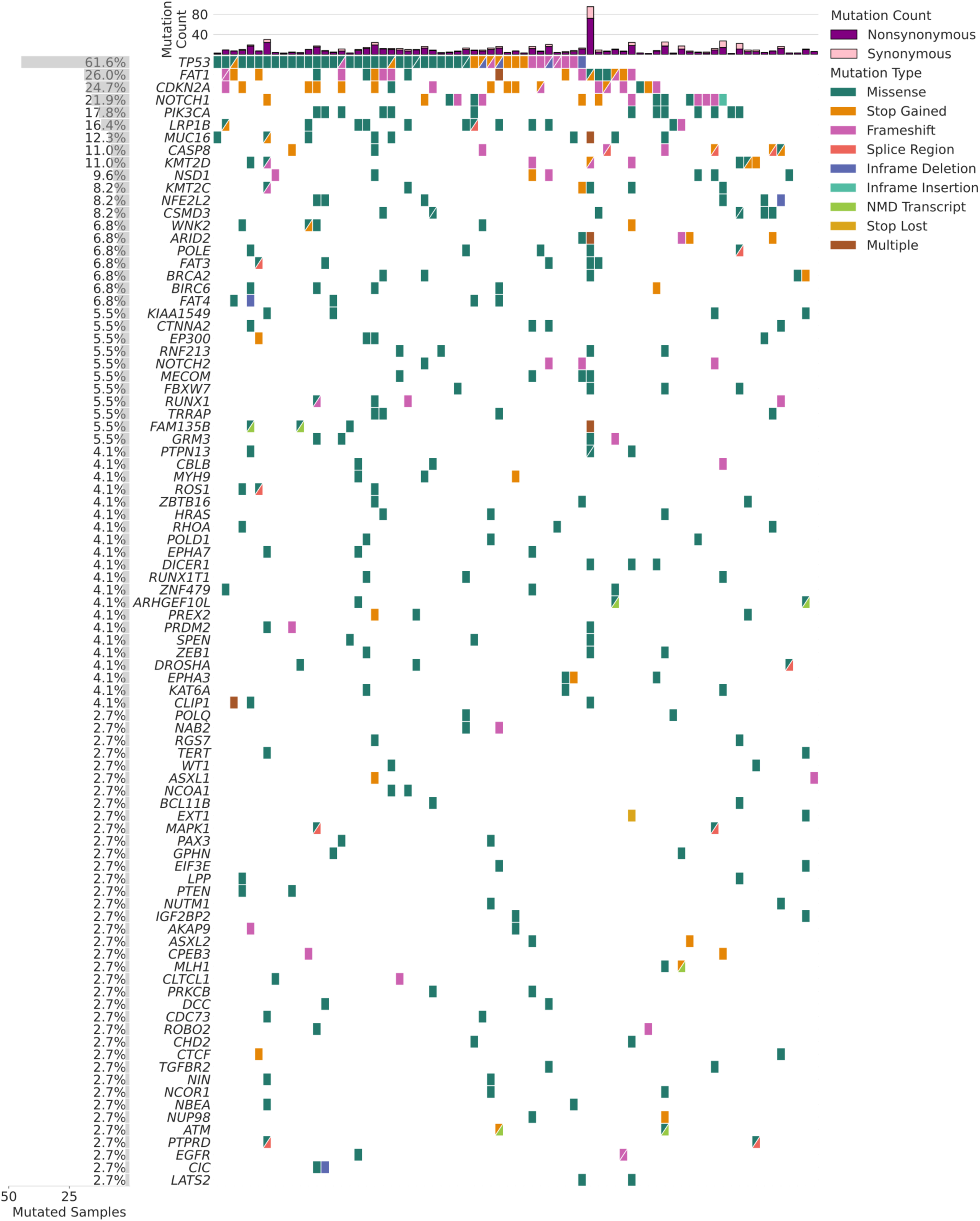
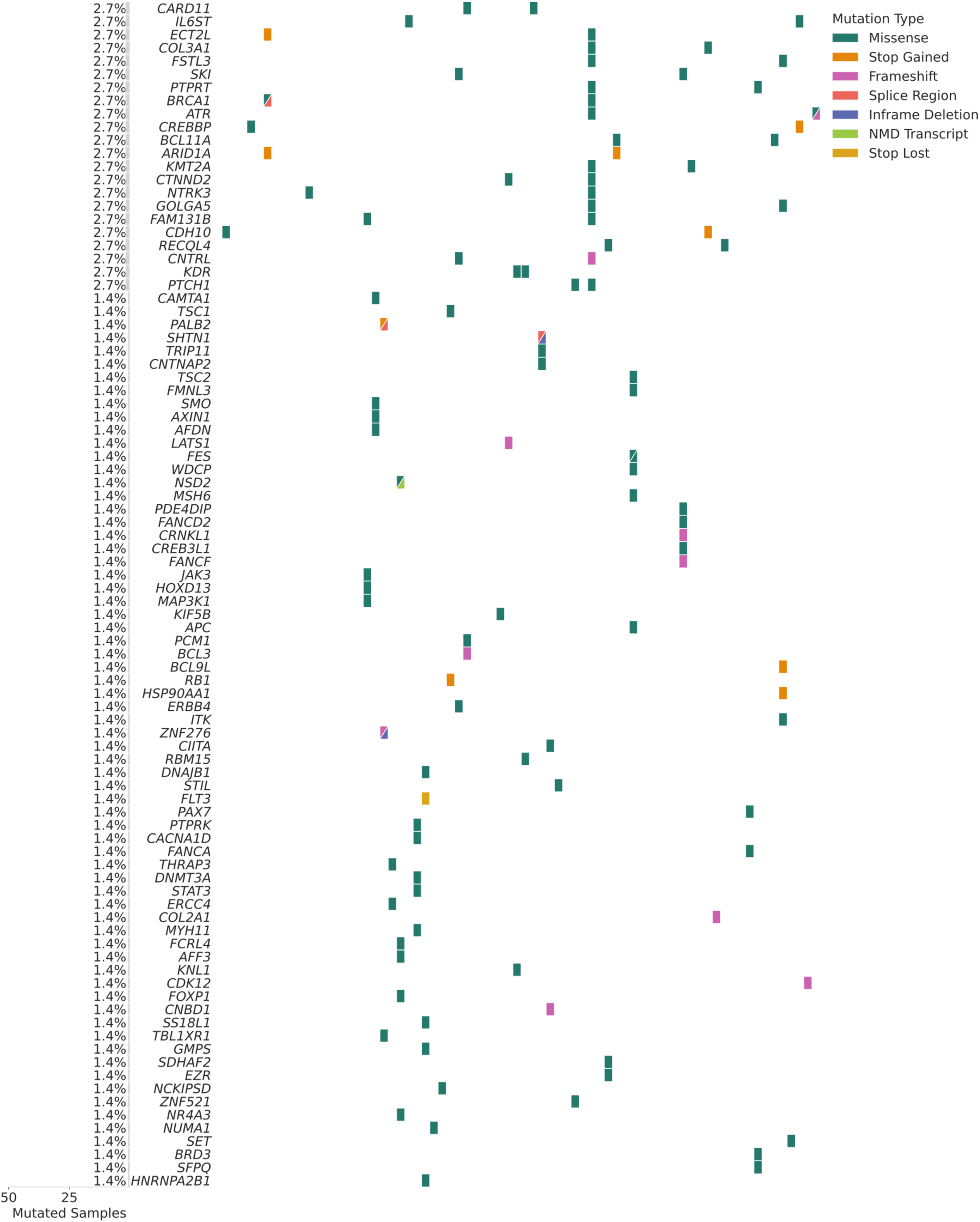
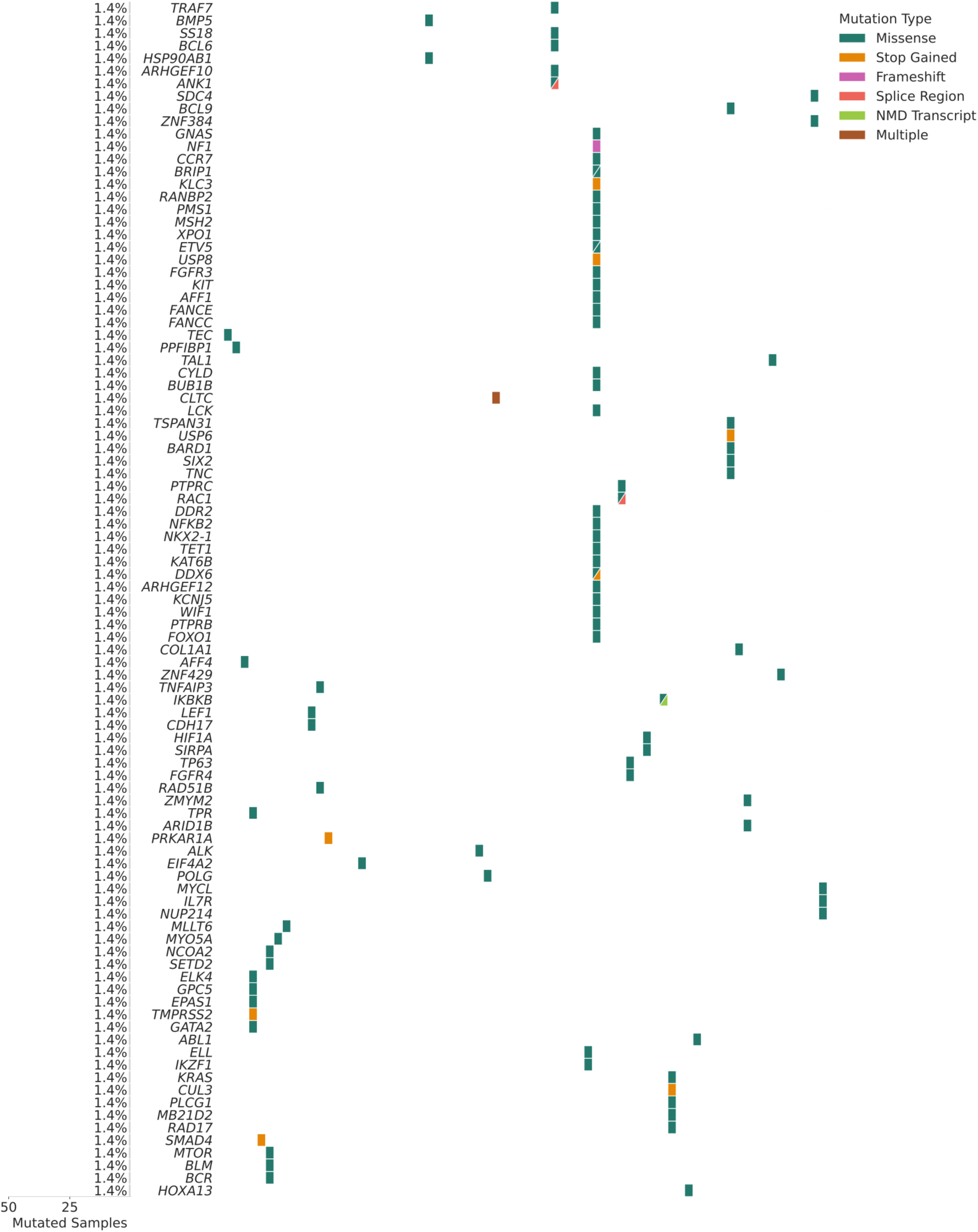
Co-mutation plot from targeted sequencing of 73 HPV-HNSCC patient tumors (all detected mutations shown).

**Supplementary Figure 3.**
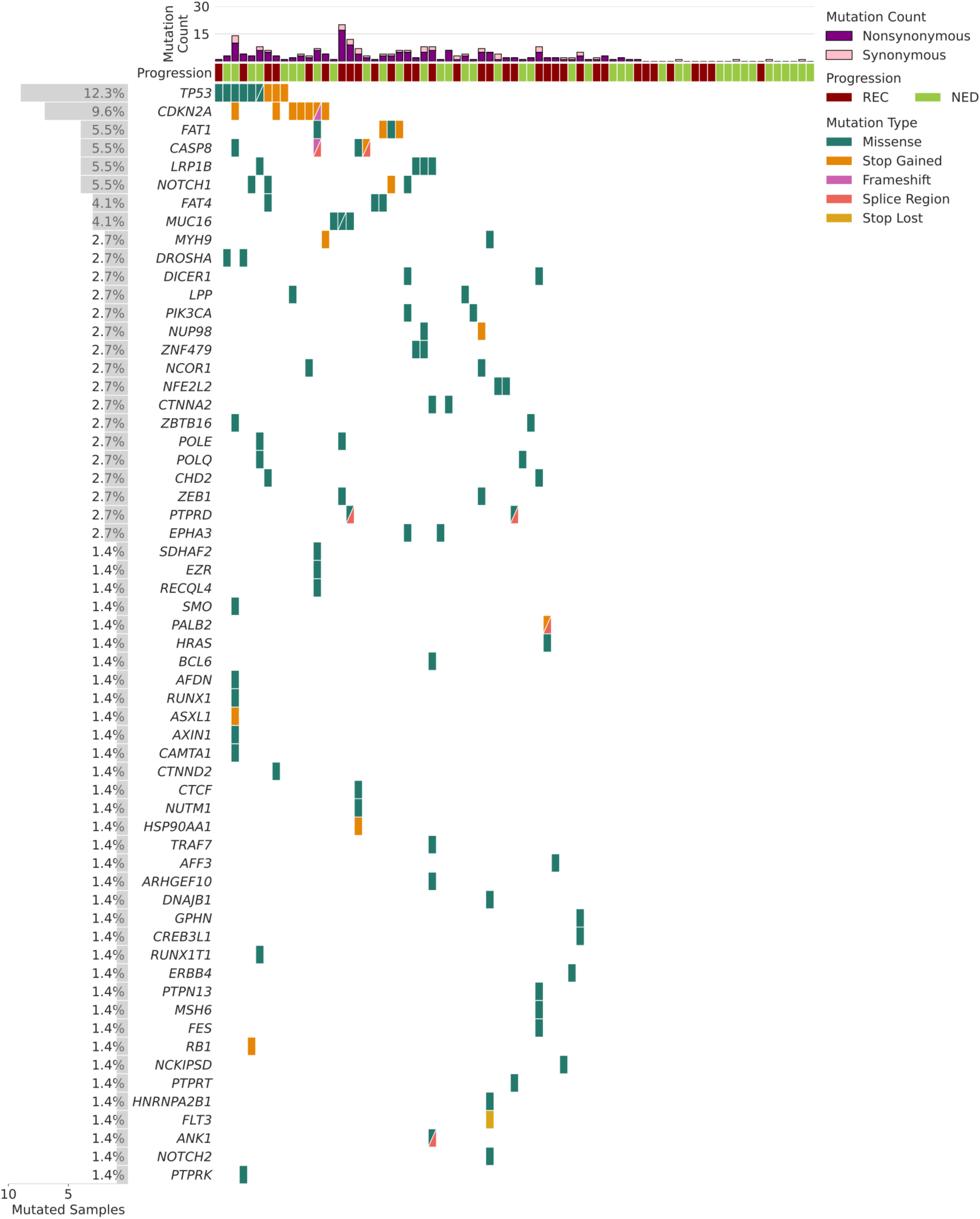
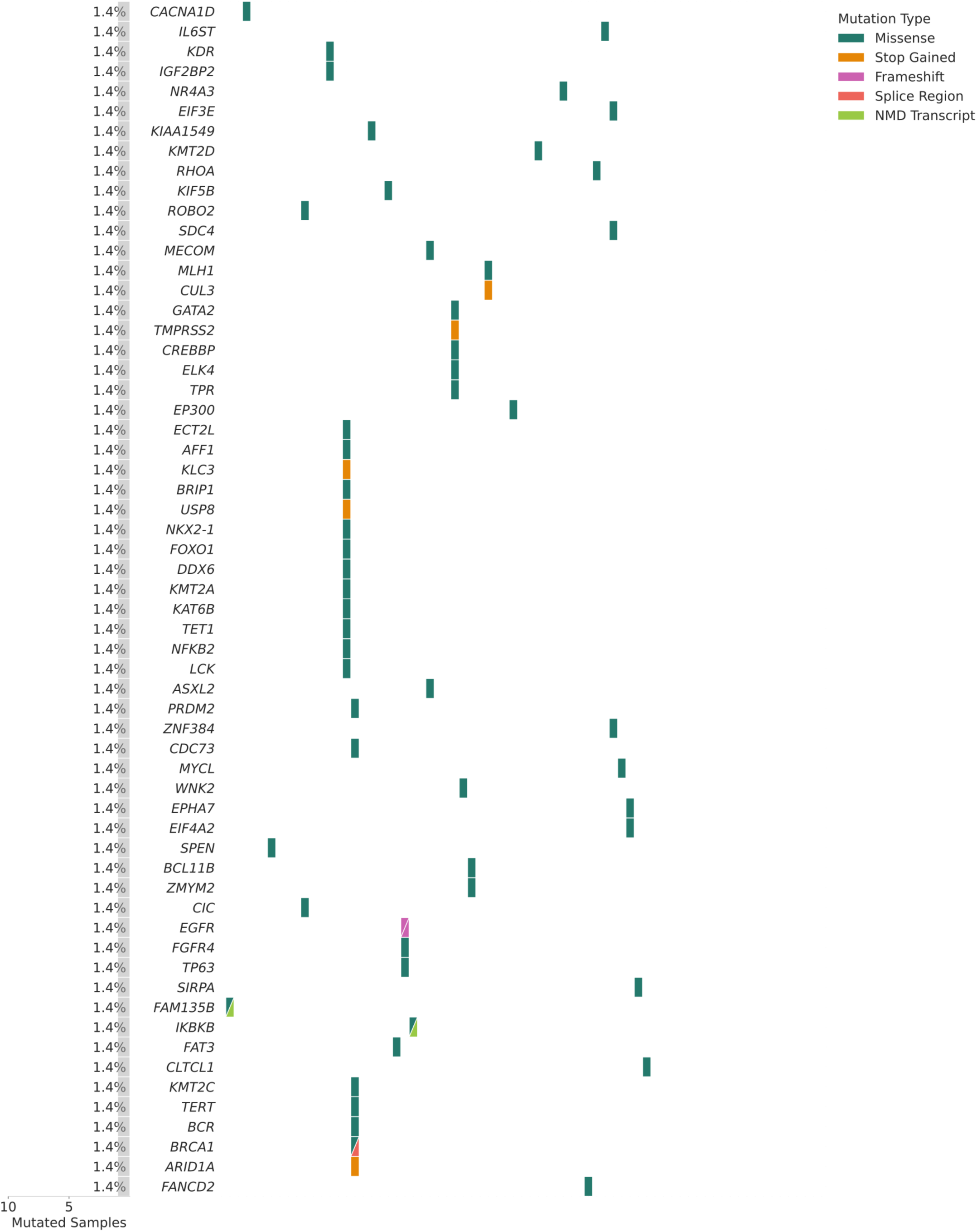
Co-mutation plot from targeted sequencing of 73 HPV-HNSCC patient lymph samples (all detected mutations shown).

**Supplementary Figure 4.**
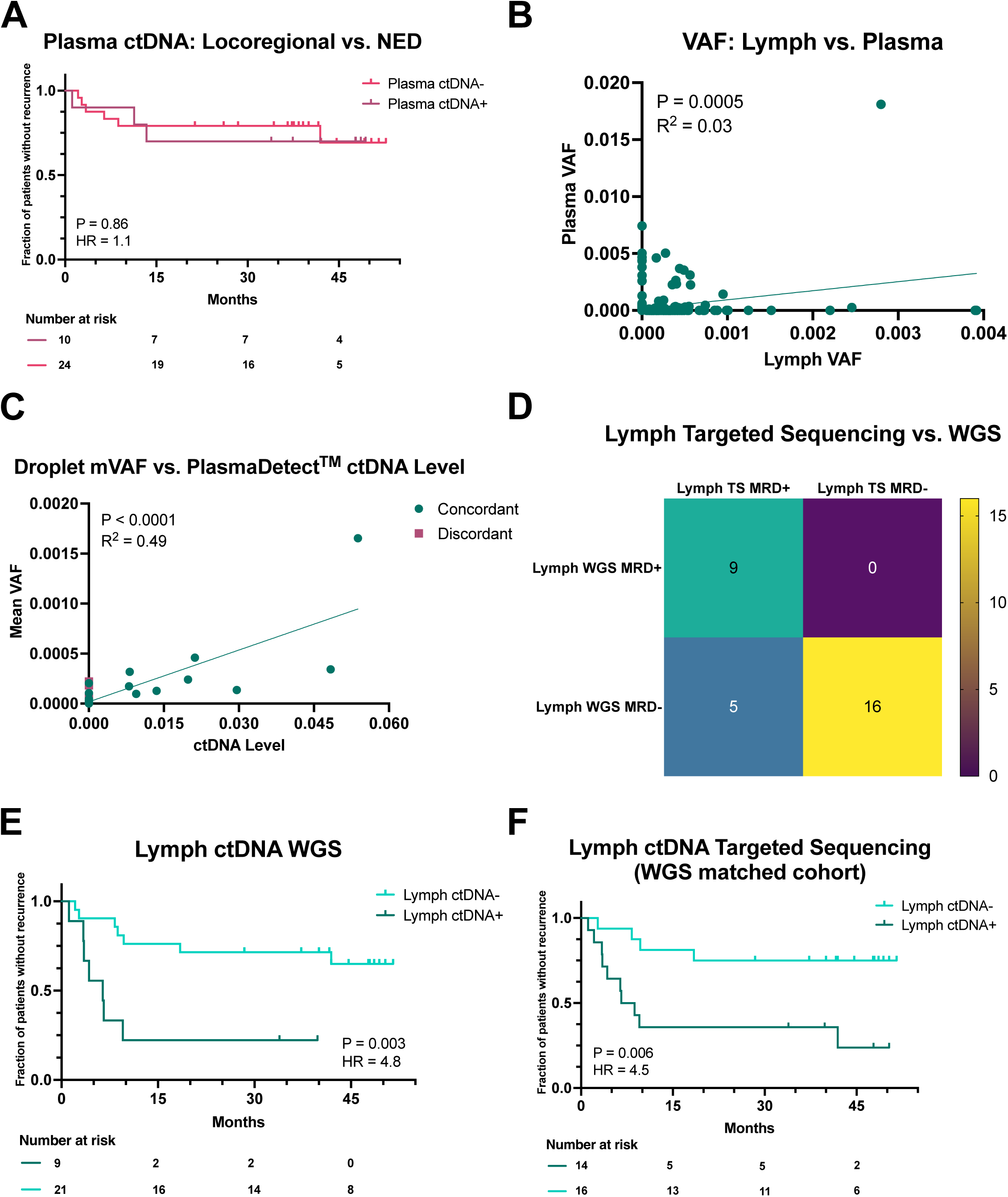
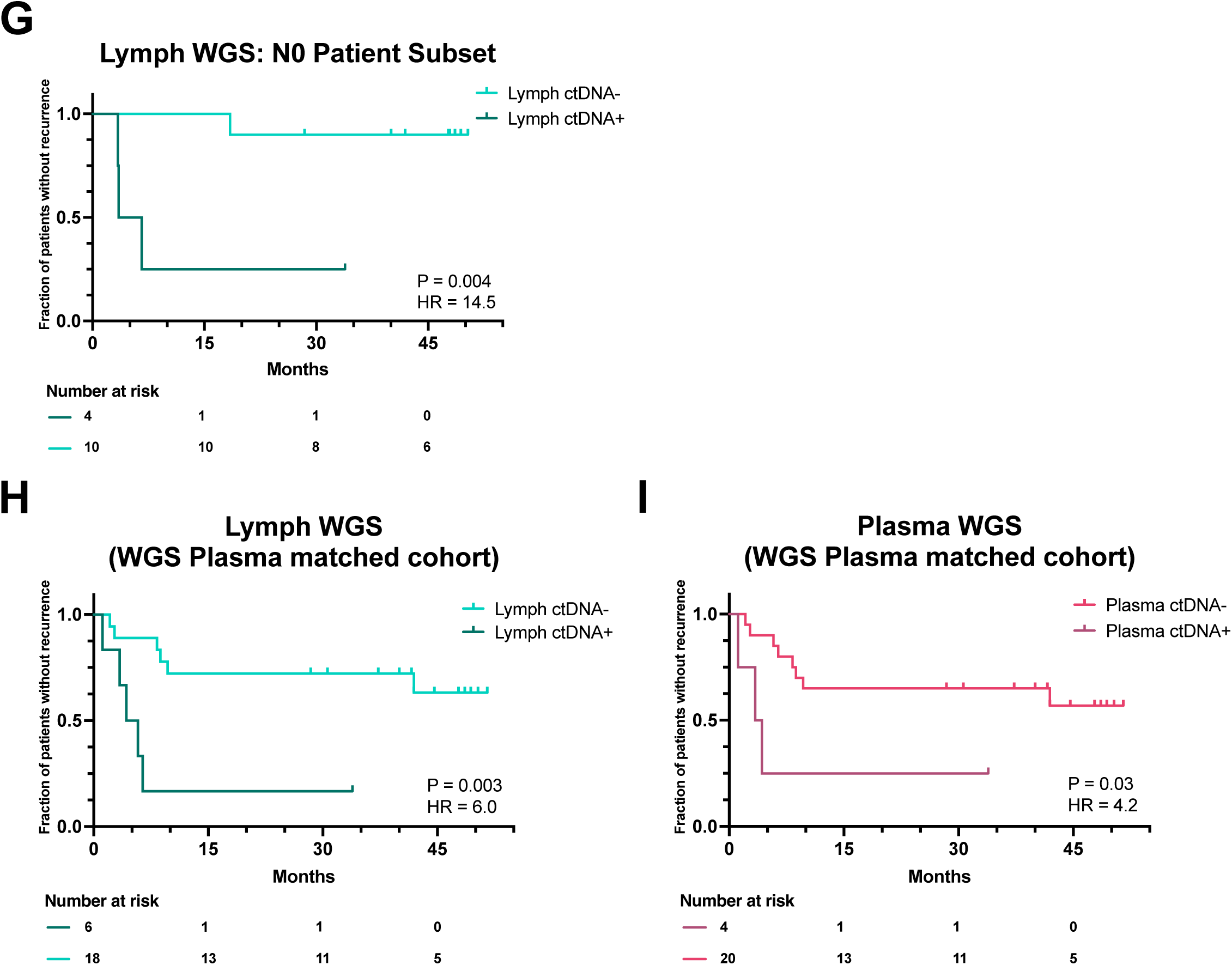
Orthogonal Validation of Lymph and Plasma MRD Detection. **A.** KM PFS analysis by Plasma ctDNA status of locoregional recurred and NED patients (n=34), sensitivity = 33%, specificity = 72%; Logrank *P* = 0.86, HR = 1.1 (95% CI, 0.3-4.6). **B.** Lymph VAF vs. Plasma VAF in matched cohort (n = 387 mutations, *P* = 0.0005, r^2^ = 0.03). **C.** Targeted sequencing mVAF vs. WGS ctDNA level (n = 30, *P* < 0.0001, r^2^ = 0.49). **D.** Targeted sequencing vs. WGS MRD prediction, *P* = 0.0001. **E.** KM PFS analysis by Lymph WGS ctDNA level (n = 30), sensitivity = 50%, specificity = 88%; *P* = 0.003, HR = 4.8 (95% CI, 1.6-14.6). **F.** KM PFS analysis by Lymph targeted sequencing ctDNA status in matched patients to WGS analysis (n = 30), sensitivity = 71%, specificity = 75%; *P* = 0.006, HR = 4.5 (95% CI, 1.4-14.5). **G.** KM PFS analysis by Lymph WGS ctDNA level in N0 patient subset (n = 14), sensitivity = 75%, specificity = 90%; *P* = 0.004, HR = 14.5 (95% CI, 1.4-148.1). **H.** KM PFS analysis by Lymph WGS ctDNA level in plasma-matched patients (n = 24), sensitivity = 45%, specificity = 92%; *P* = 0.003, HR = 6.0 (95% CI, 1.6-22.1). **I.** KM PFS analysis by Plasma WGS ctDNA level (n = 24), sensitivity = 27%, specificity = 92%, *P* = 0.03, HR = 4.2 (95% CI, 1-16.6).

**Supplementary Figure 5.**
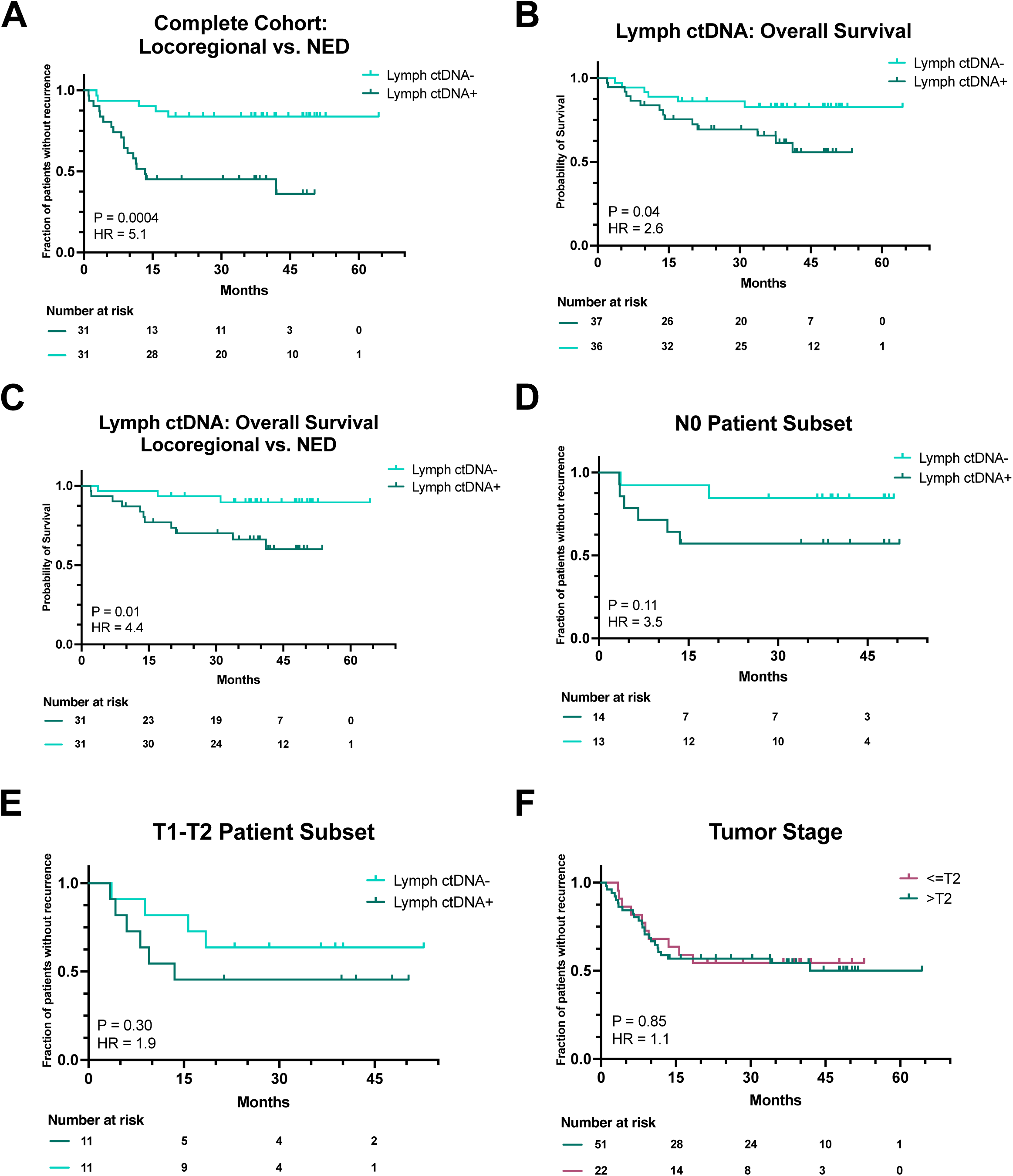
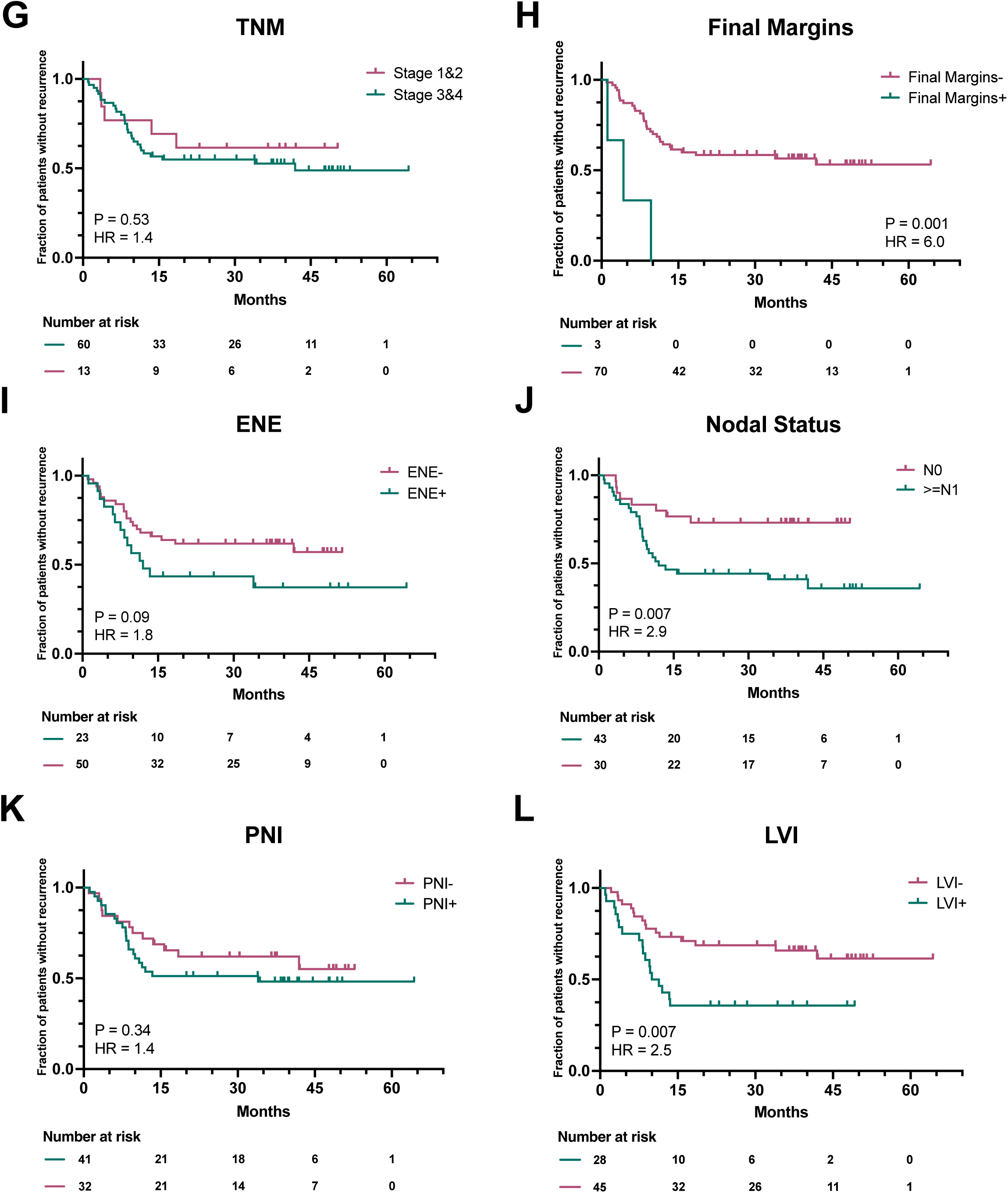
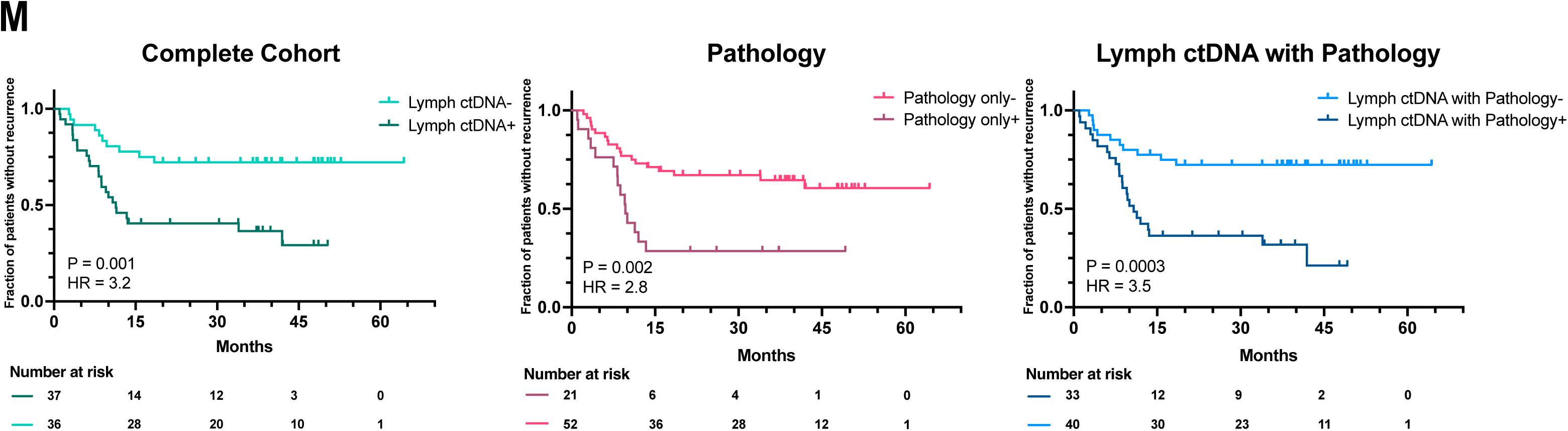
Lymph ctDNA outperforms pathology biomarkers. **A.** KM PFS analysis by Lymph ctDNA status for locoregional recurred and NED patients in Complete Cohort (n=62), sensitivity = 78%, specificity = 67%; Logrank *P* = 0.0004, HR = 5.1 (95% CI, 1.9-13.9). **B.** KM OS analysis by Lymph ctDNA status for Complete Cohort (n=73), sensitivity = 70%, specificity = 57%; Logrank *P* = 0.04, HR = 2.6 (95% CI, 1-6.8). **C.** KM OS analysis by Lymph ctDNA status for locoregional recurred and NED patients in Complete Cohort (n=62), sensitivity = 79%, specificity = 58%; Logrank *P* = 0.01, HR = 4.4 (95% CI, 1.2-15.7). **D.** KM PFS analysis by lymph ctDNA status in N0 stage patients (n=27), sensitivity =75%, specificity = 58%, Logrank *P* = 0.11, HR = 3.5 (95% CI, 0.7-17.3). **E.** KM PFS analysis by Lymph ctDNA status in T1-T2 stage patients (n=22), sensitivity = 60%, specificity = 58%, Logrank *P* = 0.3, HR = 1.9 (95% CI, 0.5-6.9). **F.** KM PFS analysis by Tumor stage (stage 3&4 vs. stage 1&2, n=73), sensitivity = 71%, specificity = 31%; Logrank *P* = 0.85, HR = 1.1 (95% CI, 0.5-2.3). **G.** KM PFS analysis by TNM system (overall stage 3&4 vs. stage 1&2, n=73), sensitivity = 85%, specificity = 21%; Logrank *P* = 0.53, HR = 1.4 (95% CI, 0.5-3.5). **H.** KM PFS analysis by Final Margins (n=73), sensitivity = 9%, specificity = 100%; Logrank *P* = 0.001, HR = 6.0 (95% CI, 1.8-20.4). **I.** KM PFS analysis by Extranodal Extension status (n=73), sensitivity = 41%, specificity = 77%; Logrank *P* = 0.09, HR = 1.8 (95% CI, 0.9-3.6). **J.** KM PFS analysis by Nodal status (N1+ vs. N0, n=73), sensitivity = 76%, specificity = 56%; Logrank *P* = 0.007, HR = 2.9 (95% CI, 1.3-6.4). **K.** KM PFS analysis by Perineural Invasion status (n=73), sensitivity = 62%, specificity = 49%; Logrank *P* = 0.34, HR = 1.4 (95% CI, 0.7-2.8). **L.** KM PFS analysis by Lymphovascular Invasion status (n=73), sensitivity = 53%, specificity = 74%; Logrank *P* = 0.007, HR = 2.5 (95% CI, 1.3-4.9). **M.** KM PFS analysis by lymph ctDNA status for Complete Cohort (n=73), sensitivity = 71%, specificity = 67%, Logrank *P* = 0.001, HR = 3.2 (95% CI, 1.5-6.8); KM PFS analysis by pathology for Complete Cohort (n = 73), sensitivity = 44%, specificity = 85%, Logrank *P* = 0.002, HR = 2.8 (95% CI, 1.4-5.6); KM PFS analysis by lymph ctDNA status + pathology for Complete Cohort (n = 73), sensitivity = 68%, specificity = 74%, Logrank *P* = 0.0003, HR = 3.5 (1.7-7.2).

